# Applying machine-learning to rapidly analyse large qualitative text datasets to inform the COVID-19 pandemic response: Comparing human and machine-assisted topic analysis techniques

**DOI:** 10.1101/2022.05.12.22274993

**Authors:** Lauren Towler, Paulina Bondaronek, Trisevgeni Papakonstantinou, Richard Amlôt, Tim Chadborn, Ben Ainsworth, Lucy Yardley

## Abstract

**Background:** Machine-assisted topic analysis (MATA) uses artificial intelligence methods to assist qualitative researchers to analyse large amounts of textual data. This could allow qualitative researchers to inform and update public health interventions ‘in real-time’, to ensure they remain acceptable and effective during rapidly changing contexts (such as a pandemic). In this novel study we aimed to understand the potential for such approaches to support intervention implementation, by directly comparing MATA and ‘human-only’ thematic analysis techniques when applied to the same dataset (1472 free-text responses from users of the COVID-19 infection control intervention ‘Germ Defence’).

**Methods:** In MATA, the analysis process included an unsupervised topic modelling approach to identify latent topics in the text. The human research team then described the topics and identified broad themes. In human-only codebook analysis, an initial codebook was developed by an experienced qualitative researcher and applied to the dataset by a well-trained research team, who met regularly to critique and refine the codes. To understand similarities and difference, formal triangulation using a ‘convergence coding matrix’ compared the findings from both methods, categorising them as ‘agreement’, ‘complementary’, ‘dissonant’, or ‘silent’.

**Results:** Human analysis took much longer (147.5 hours) than MATA (40 hours). Both human-only and MATA identified key themes about what users found helpful and unhelpful (e.g. *Boosting confidence in how to perform the behaviours* vs *Lack of personally relevant content*). Formal triangulation of the codes created showed high similarity between the findings. All codes developed from the MATA were classified as in agreement or complementary to the human themes. Where the findings were classified as complementary, this was typically due to slightly differing interpretations or nuance present in the human-only analysis.

**Conclusions:** Overall, the quality of MATA was as high as the human-only thematic analysis, with substantial time savings. For simple analyses that do not require an in-depth or subtle understanding of the data, MATA is a useful tool that can support qualitative researchers to interpret and analyse large datasets quickly. These findings have practical implications for intervention development and implementation, such as enabling rapid optimisation during public health emergencies.

**Contributions to the literature:**

- Natural language processing (NLP) techniques have been applied within health research due to the need to rapidly analyse large samples of qualitative data. However, the extent to which these techniques lead to results comparable to human coding requires further assessment.
- We demonstrate that combining NLP with human analysis to analyse free-text data can be a trustworthy and efficient method to use on large quantities of qualitative data.
- This method has the potential to play an important role in contexts where rapid descriptive or exploratory analysis of very large datasets is required, such as during a public health emergency.

## Background

Qualitative research plays a vital role in public health, intervention development and implementation research by enabling researchers to develop an informed understanding of the attitudes, perceptions and contextual factors relevant to planning and delivering effective and acceptable health interventions (Hamilton & Finley, 2019; Shuval et al., 2011). However, most qualitative approaches (such as interviews, focus groups and observation studies) are resource intensive and time-consuming, requiring months or years to collect and analyse rich, in-depth data. Consequently, most qualitative approaches have traditionally been based on studies of relatively small, purposively selected samples (Vindrola-Padros et al., 2020). While this kind of in-depth approach has enormous benefits in terms of generating nuanced insights for the purpose of theory-building, it is less suitable for some potential applications of qualitative methods. In particular, less resource intensive methods are needed in order to analyse the wealth of qualitative data that can be generated by automated online data collection (for example, of free text responses to population surveys).

Recent advances in technology have facilitated the automatic processing of text-based qualitative datasets, via natural language processing (NLP), a subfield of artificial intelligence. NLP algorithms can quickly produce ‘triaged’ natural text outputs, that have the potential to substantially reduce the amount of text to be examined by research teams while remaining meaningful (Crowston et al., 2012). NLP has been applied in several areas of healthcare research: extracting information from electronic healthcare records (Ford et al., 2016; Zheng et al., 2016), coding interview transcripts about male health needs (Leeson et al., 2019), or early detection of depression in social networks (Cacheda et al., 2019). A direct comparison of an NLP approach which used lexicon-based clustering in WordNet with human-only qualitative analysis analysed answers from 84 participants to short open-ended text message survey questions (Guetterman et al., 2018). They found that NLP generated similar findings although was not of as high quality, and could be used to in combination with human qualitative analysis to provide more detail.

Indeed, the importance of the input of experienced qualitative researchers to NLP-assisted qualitative data analysis must not be overlooked. Findings by Guetterman and colleagues (2018) highlight how experienced qualitative researchers bring knowledge of contextual, theoretical, and sociocultural factors that cannot be replicated by NLP-only approaches. While previous studies show how NLP methods can be used to support deductive approaches where an a priori coding framework is in place (Lennon et al., 2021), there is often a need to conduct ‘bottom-up’ inductive and exploratory analyses where ideas are formed from the data itself, particularly when developing new public health interventions or adapting existing interventions to new situations or populations. Inductive qualitative analysis allows researchers to explore relevant issues and topics as guided by members of the relevant population, and generate new ideas in a data-driven way (Braun & Clarke, 2013; Greenhalgh & Taylor, 1997). In this project, we therefore aimed to explore the use of a different specific NLP approach which integrates human and exploratory NLP analysis– which we have termed “Machine-Assisted Topic Analysis” (MATA) – to allow expert qualitative researchers to look at large, real-world datasets in a timely manner.⍰

MATA assists qualitative researchers by summarising major patterns in the text according to generative models of word counts – known as topic models (Roberts et al., 2019). Topic models are able to automatically infer latent topics from text. This means the model assumes that the documents consist of a combination of underlying topics and can be represented as such. Topic models allow for machine-assisted reading of text datasets through creating and extracting the main themes that underlie a corpus and mapping them onto the individual documents. They are particularly useful as tools to analyse large volumes of free-text responses to questions in a data-driven way, in order to summarise the main families of responses. The approach used in this study is based on an application of the Structural Topic Model (Roberts et al., 2013; 2019) in particular. The STM is a general framework for topic modelling that is differentiated from other topic modelling methodologies by its ability to enable researchers to include additional variables at the document level, such as the date a document was created or the demographics of the person who created it, as covariates in a topic model. This way the relationships of these variables to specific topics can be estimated and examined or used to run subgroup analyses. Those variables are further used to explain variance in topic prevalence, so affect the frequency with which a topic is discussed. As a result, their inclusion improves inference and qualitative interpretability and also affects the topical content (Roberts et al., 2018). Structural topic models are able to identify patterns, and qualitative researchers can then use the output to extract meaning, interpret and summarise the topics.

Within the context of COVID-19, several NLP researchers have identified NLP as a potentially effective tool for rapid analysis of large-scale text-based datasets in order to meet the rapidly shifting public health needs during a pandemic (Baclic et al., 2020; Chang et al., 2020, Lennon et al., 2021). For example, NLP approaches could allow the rapid analysis of views and experiences of public health interventions (such as infection tracking tools, or public health messaging services) via survey response, allowing teams to improve interventions in real-time as issues arise – which can be vital given the rapidly changing context of a worldwide pandemic (Morton et al., 2021; Vindrola-Padros et al., 2020). However, previous comparisons between exploratory NLP methods and human-only qualitative analyses have mostly been conducted on relatively small sample sizes (Guetterman et al., 2018; Leeson et al., 2019). Therefore, there is a need to assess how NLP methods can inductively analyse large datasets for studies with exploratory aims.

Germ Defence is a digital behaviour change intervention that aims to improve infection control behaviours during the COVID-19 pandemic (Ainsworth et al., 2021). In order to remain as effective as possible, Germ Defence was iteratively updated throughout the pandemic, as health guidelines and contextual factors (e.g. virus prevalence, vaccine uptake) changed (Morton et al., 2021). During the intervention, some website users provided feedback about the content and design, and we used this data to perform separate qualitative analyses using MATA and human-only analysis. We aimed to explore similarities and differences between findings of the two methods, and to compare the person-hours required to conduct each form of analysis, in order to assess the potential value and trustworthiness of MATA for large-scale public health intervention evaluation and optimisation.

## Methods

### Participants

Inclusion criteria were users of the Germ Defence website who were over the age of 18 and able to give informed consent. Between 18th November 2020 until 3rd January 2021, a total of 2175 people consented to the survey, 1472 of which responded to at least one open-ended question. During this time, a second national lockdown was in place in the UK, which was replaced by the reintroduction of the tiered system on 2nd December 2020. Data collection ended prior to the third national lockdown on 6th January 2021. Table 1 shows the demographic characteristics of the sample.

**Table 1.**
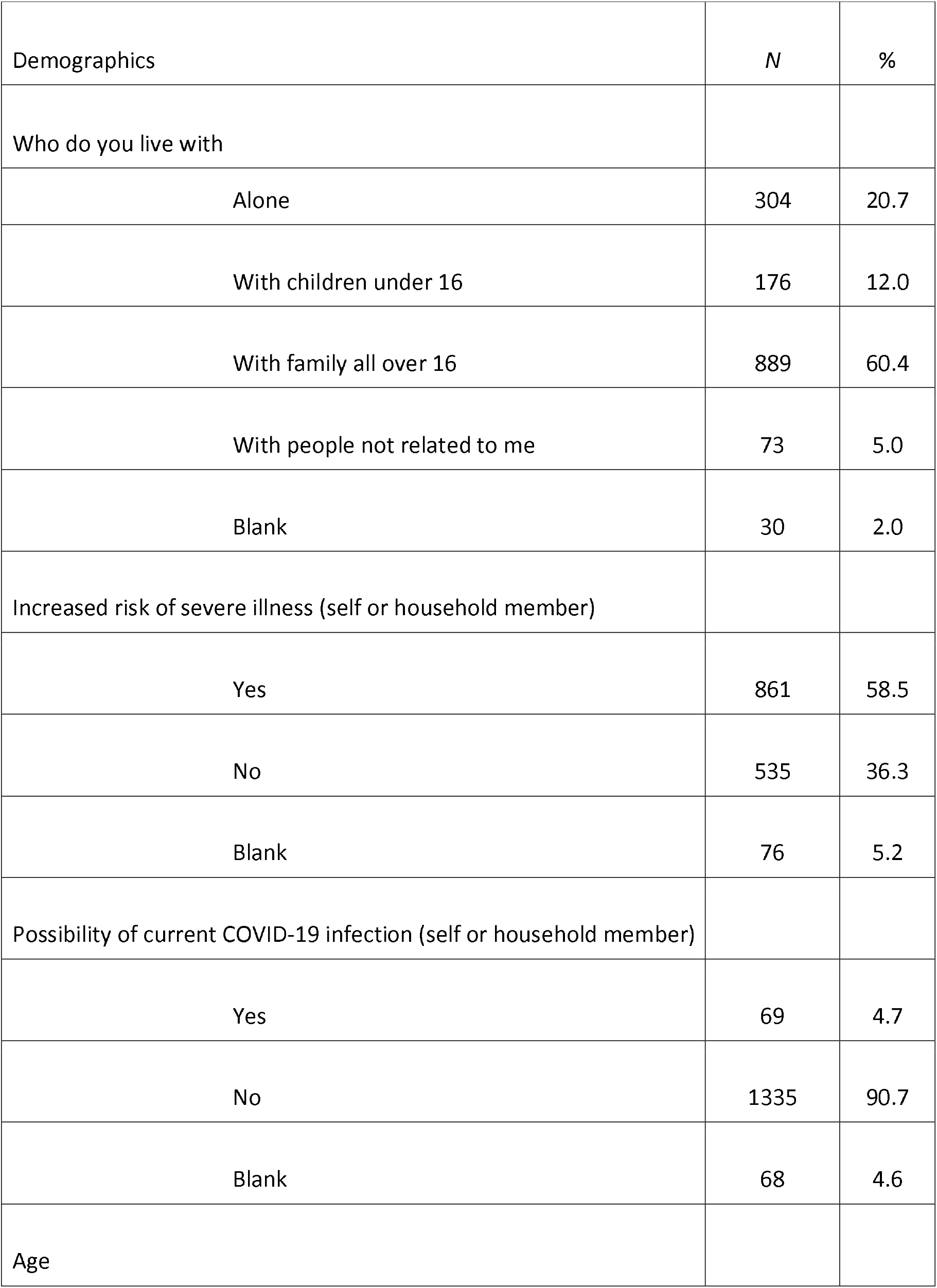
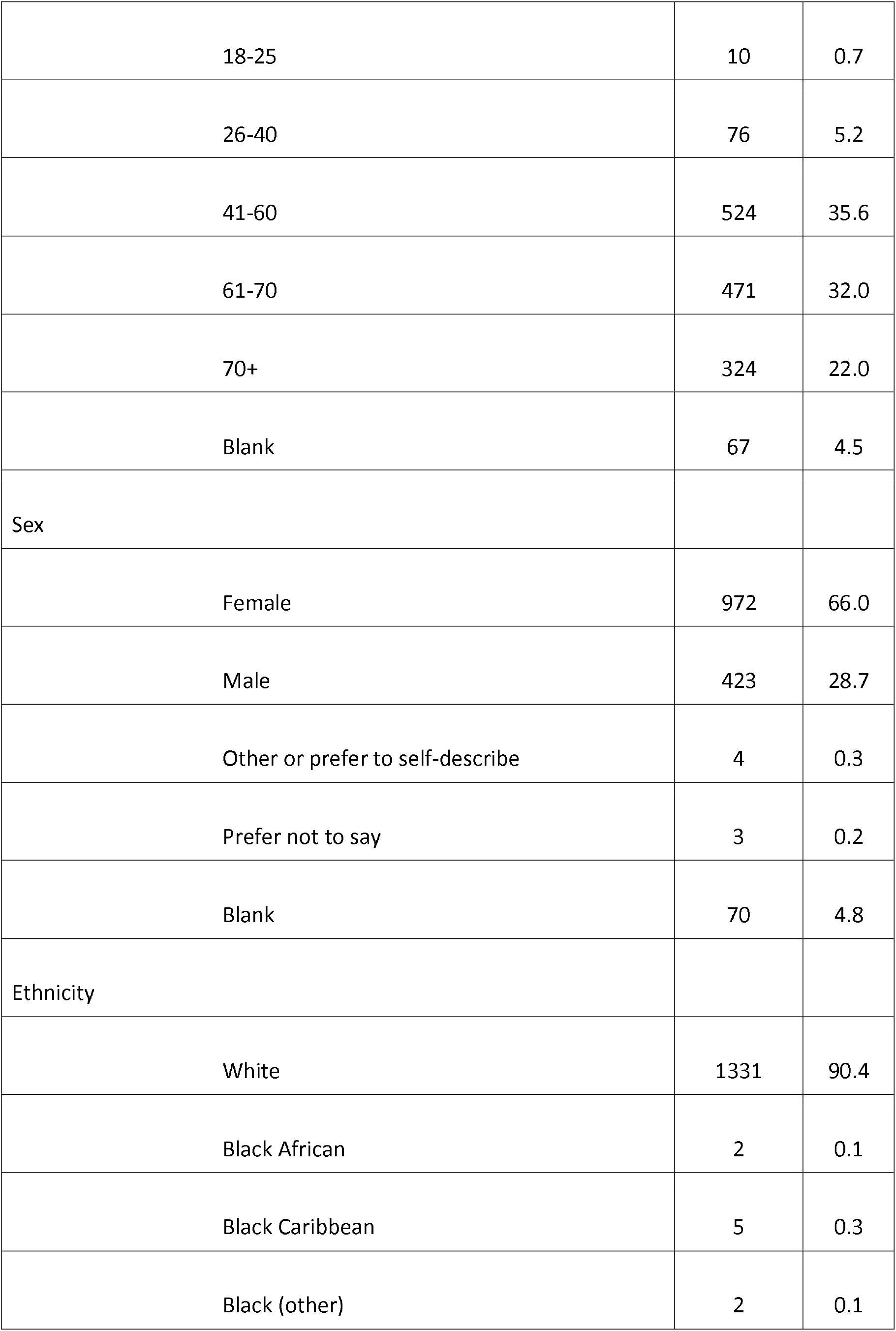
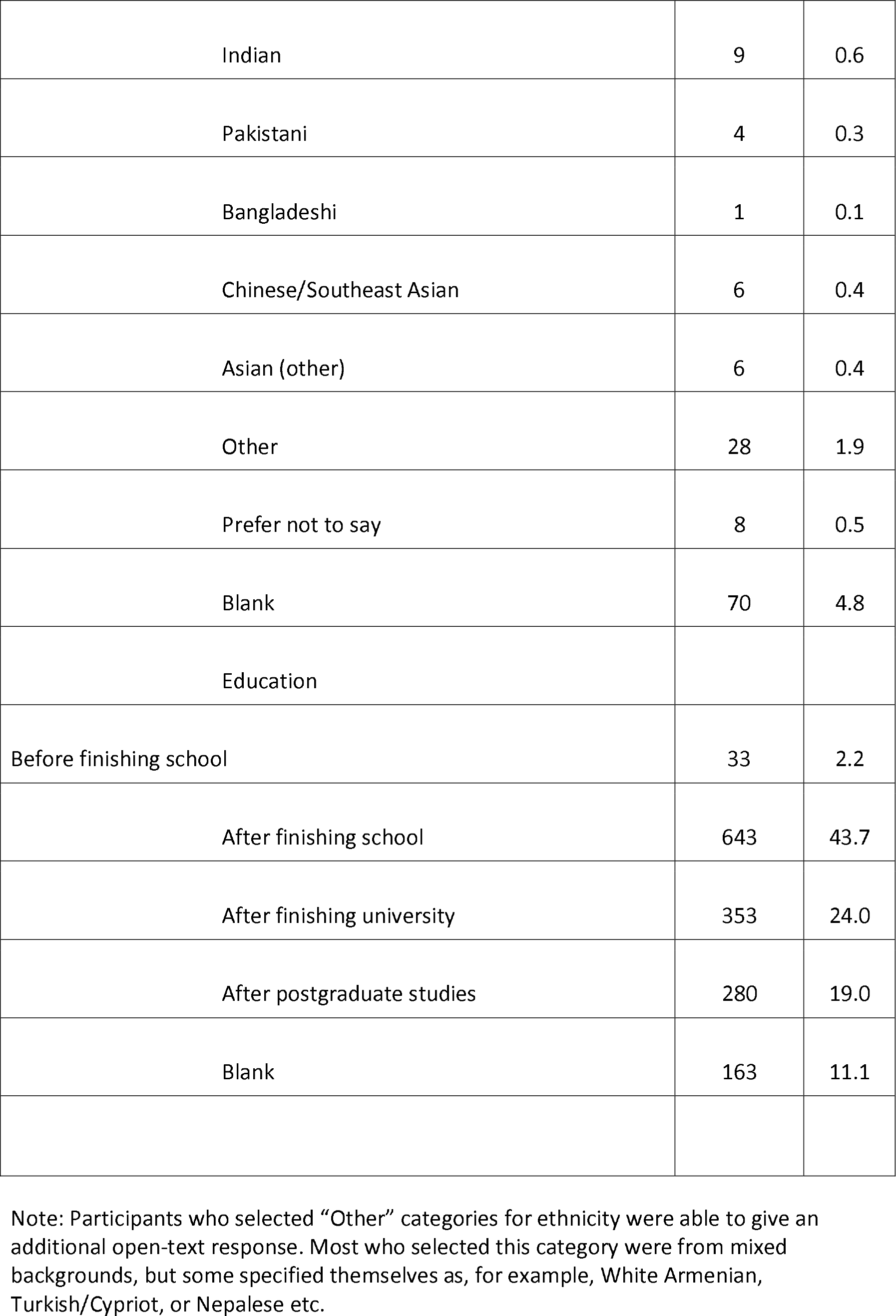
Demographic characteristics of the sample (*N*=1472)

| Demographics | *N* | % |
| --- | --- | --- |
| Who do you live with |  |  |
| Alone | 304 | 20.7 |
| With children under 16 | 176 | 12.0 |
| With family all over 16 | 889 | 60.4 |
| With people not related to me | 73 | 5.0 |
| Blank | 30 | 2.0 |
| Increased risk of severe illness (self or household member) |  |  |
| Yes | 861 | 58.5 |
| No | 535 | 36.3 |
| Blank | 76 | 5.2 |
| Possibility of current COVID-19 infection (self or household member) |  |  |
| Yes | 69 | 4.7 |
| No | 1335 | 90.7 |
| Blank | 68 | 4.6 |
| Age |  |  |
| 18-25 | 10 | 0.7 |
| 26-40 | 76 | 5.2 |
| 41-60 | 524 | 35.6 |
| 61-70 | 471 | 32.0 |
| 70+ | 324 | 22.0 |
| Blank | 67 | 4.5 |
| Sex |  |  |
| Female | 972 | 66.0 |
| Male | 423 | 28.7 |
| Other or prefer to self-describe | 4 | 0.3 |
| Prefer not to say | 3 | 0.2 |
| Blank | 70 | 4.8 |
| Ethnicity |  |  |
| White | 1331 | 90.4 |
| Black African | 2 | 0.1 |
| Black Caribbean | 5 | 0.3 |
| Black (other) | 2 | 0.1 |
| Indian | 9 | 0.6 |
| Pakistani | 4 | 0.3 |
| Bangladeshi | 1 | 0.1 |
| Chinese/Southeast Asian | 6 | 0.4 |
| Asian (other) | 6 | 0.4 |
| Other | 28 | 1.9 |
| Prefer not to say | 8 | 0.5 |
| Blank | 70 | 4.8 |
| Education |  |  |
| Before finishing school | 33 | 2.2 |
| After finishing school | 643 | 43.7 |
| After finishing university | 353 | 24.0 |
| After postgraduate studies | 280 | 19.0 |
| Blank | 163 | 11.1 |

Note: Participants who selected “Other” categories for ethnicity were able to give an additional open-text response. Most who selected this category were from mixed backgrounds, but some specified themselves as, for example, White Armenian, Turkish/Cypriot, or Nepalese etc.

### Measures

To gather demographic data, closed questions were asked pertaining to age, sex, ethnicity, education, household size, whether the user or someone else in the household is at increased risk of severe illness if they caught COVID, and whether there could be a current COVID case within the household (experiencing symptoms or contact with confirmed case). Feedback was collected as free-text responses to two questions: “What was helpful about the information on the Germ Defence website?” and “What did you not find helpful about the information on the Germ Defence website?” Responses to these questions provide a rich dataset of recommendations that can be used to improve the website and guidance provided.

### Procedure

After they had completed at least one of the two main sections of the intervention (handwashing or reducing illness), visitors to the Germ Defence website received a pop-up asking if they might be interested in taking a survey to help improve the website. The invitation was presented as seeking information on users’ views on protecting themselves from Coronavirus, and their thoughts on the Germ Defence website. Users could then follow a link to the study information sheet, consent form, and the online questionnaire hosted on Qualtrics. Ethical approval was granted by the University of Southampton Psychology Ethics Committee (ID: 56445).

### Data analysis

We analysed the data in two ways; human-only qualitative analysis and MATA. The human-only analysis was conducted using a codebook thematic analysis (TA) approach using template analysis techniques (Braun & Clarke, 2019, 2021; Brooks et al., 2015) whereby the coding template was applied to the data by several coders, and the unit of analysis was free-text participant response. The initial codebook had been developed through the researchers’ (LT) contextual knowledge, involvement in collating feedback for the person-based approach (PBA) development of the Germ Defence intervention, and derived from smaller-scale survey data and formal TA of qualitative interviews with website users (Morton et al., 2021). Any proposed additional inductive codes identified during coding were discussed with the group as soon as possible, so that each coder could keep it in mind for their own coding. See Table 2 for further information on how the codebook was developed, and the procedures used in the human analysis. In the MATA, we also used template analysis techniques to analyse the topics generated by the STM, with each topic being the unit of analysis.

**Table 2.**
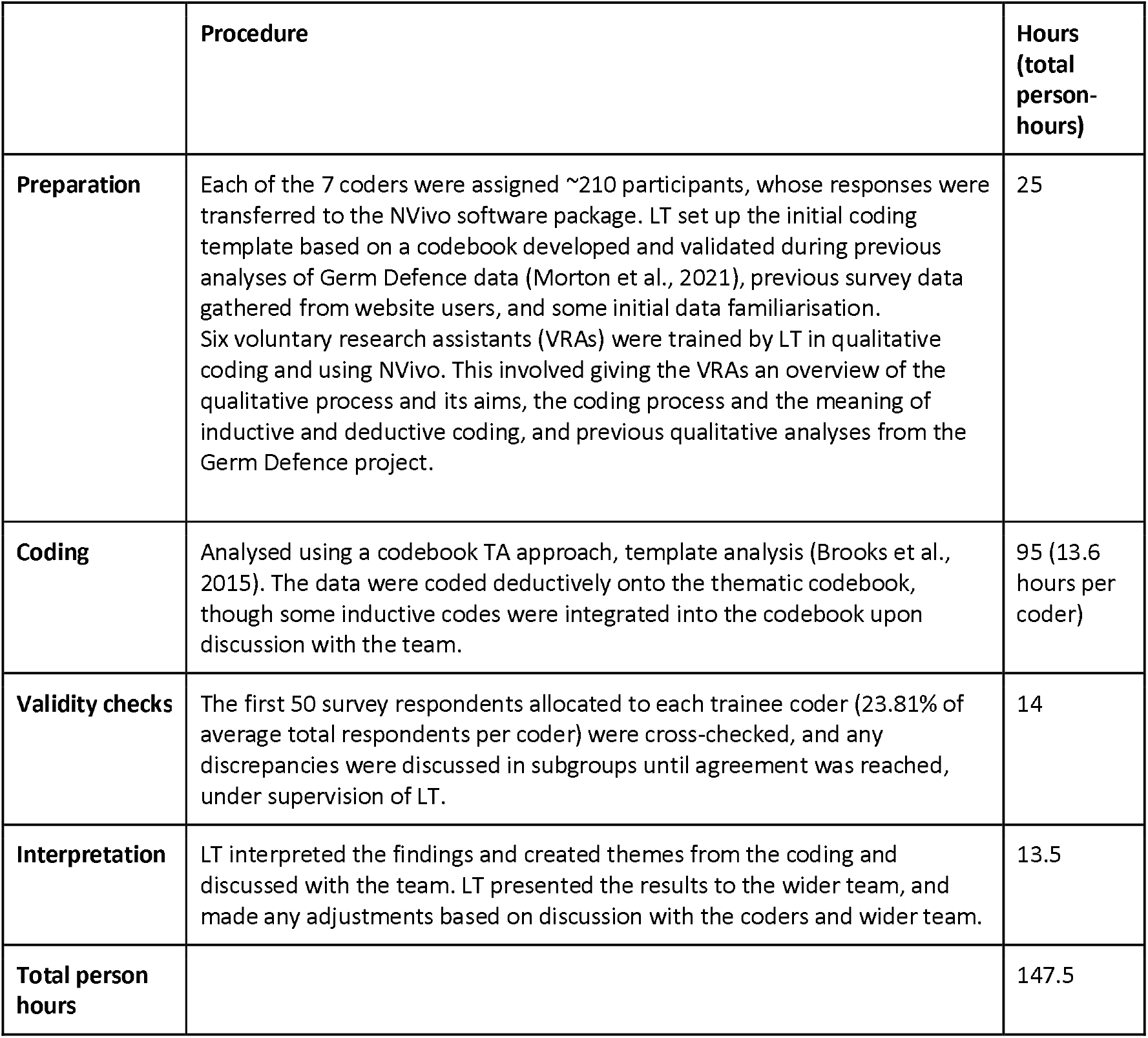
Human-only analysis procedure and person-hours

|  | **Procedure** | **Hours**<br>**(total person-hours)** |
| --- | --- | --- |
| **Preparation** | Each of the 7 coders were assigned ~210 participants, whose responses were transferred to the NVivo software package. LT set up the initial coding template based on a codebook developed and validated during previous analyses of Germ Defence data (Morton et al., 2021), previous survey data gathered from website users, and some initial data familiarisation. Six voluntary research assistants (VRAs) were trained by LT in qualitative coding and using NVivo. This involved giving the VRAs an overview of the qualitative process and its aims, the coding process and the meaning of inductive and deductive coding, and previous qualitative analyses from the Germ Defence project. | 25 |
| **Coding** | Analysed using a codebook TA approach, template analysis (Brooks et al., 2015). The data were coded deductively onto the thematic codebook, though some inductive codes were integrated into the codebook upon discussion with the team. | 95 (13.6 hours per coder) |
| **Validity checks** | The first 50 survey respondents allocated to each trainee coder (23.81% of average total respondents per coder) were cross-checked, and any discrepancies were discussed in subgroups until agreement was reached, under supervision of LT. | 14 |
| **Interpretation** | LT interpreted the findings and created themes from the coding and discussed with the team. LT presented the results to the wider team, and made any adjustments based on discussion with the coders and wider team. | 13.5 |
| **Total person hours** |  | 147.5 |

#### Machine-Assisted Topic Analysis (MATA)

##### Data

Structured data, such as date, age, sex, education level and ethnicity, were also collected and included in the models as covariates.

###### 1.1. Preparation

We preprocessed the data using R (version 3.5.2), and cleaned the free text responses using base R functions, the quanteda (version 2.0.1; Benoit et al., 2018) and stm (version 1.3.3; Roberts et al., 2019) packages. We deleted observations with missing values and duplicate data. The free-text responses were converted into token units using the quanteda package, after punctuation, symbols and numbers were removed. In this instance the tokens were individual words. Data pre-processing was completed by deleting stop words and stemming the tokens. Stemming is the process of reducing words to their root. This acts as a normalisation of text data and helps reduce the size of the dictionary which speeds up processing.

###### 1.2. Coding and validity checks

Prior to running the models we ran diagnostics to identify the optimal number of topics, according to both the relevant metrics and the aims of the analysis, focusing on the trade-off between semantic coherence and exclusivity (see Roberts et al., 2014 for a discussion on this method of evaluation). We evaluated an unsupervised Topic Modelling approach, testing models with 5-40 topics and differing covariates in terms of coherence, residuals and interpretability by human coders (see online supplementary material 1), separately for each question. Upon visually examining the plots in online supplementary material 2, we identified a Structural Topic Model with 25 topics to be optimal for addressing question A, “What was helpful about the information on the Germ Defence website?” whereas 15 topics were deemed to be optimal for addressing question B, “What did you not find helpful about the information on the Germ Defence website?”. In both cases date, age, gender, ethnicity, and level of education were included as covariates. The model automated the equivalent of the coding stage of the analysis by assigning a number of labels to each document, by way of mapping them to topics.

##### 2. Interpretation: qualitative analysis of machine-generated data by trained, supervised coders

The outputs examined consisted of two main elements; the 10 most representative quotes for each topic and two lists of weighted words that constitute the topic. Different types of word weightings were generated with each topic where the following two types were analysed in subsequent qualitative analysis: 1) Highest Prob (words within each topic with the highest probability) and 2) FREX (words that are both frequent and exclusive, identifying words that distinguish topics). Examples of outputs generated are presented in Box 1 and 2 in online supplementary material 3.

In order to analyse the model’s output systematically we analysed it in two stages. In Stage 1, two researchers interpreted the output and agreed upon narrative labels for the topics (henceforth, MATA codes). In Stage 2, the researchers analysed the topics generated by the text analysis and created broader themes. Table 3 provides a breakdown of the steps of the MATA process, along with the person-hours that were spent on each step.

**Table 3.**
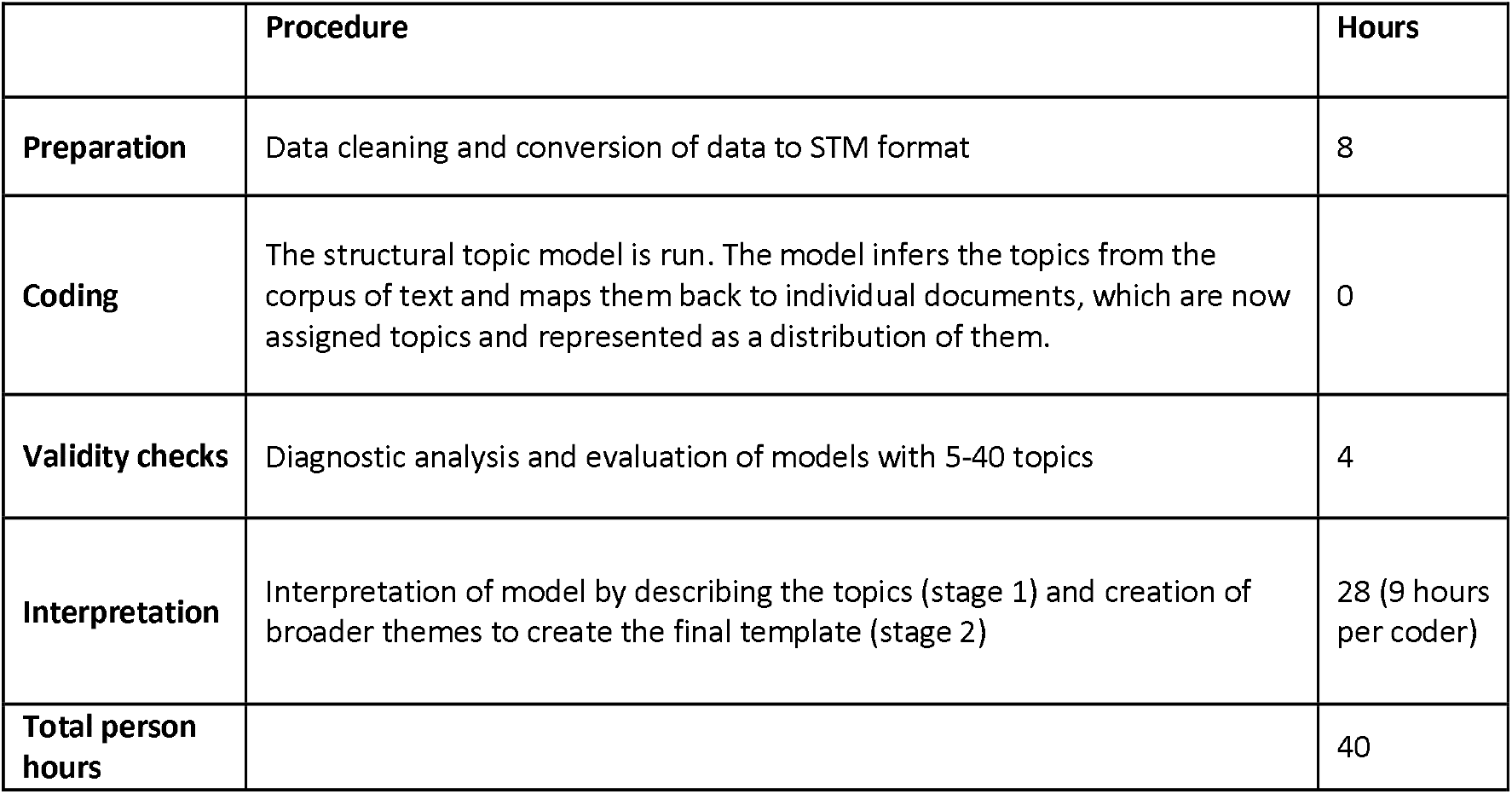
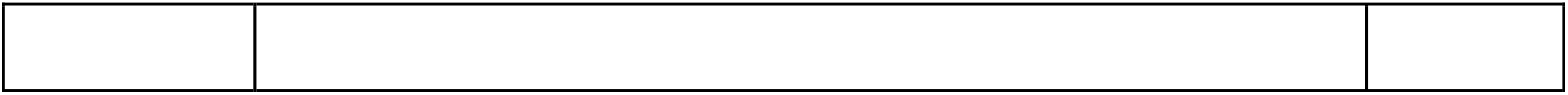
Machine-assisted topic analysis approach and person-hours

|  | **Procedure** | **Hours** |
| --- | --- | --- |
| **Preparation** | Data cleaning and conversion of data to STM format | 8 |
| **Coding** | The structural topic model is run. The model infers the topics from the corpus of text and maps them back to individual documents, which are now assigned topics and represented as a distribution of them. | 0 |
| **Validity checks** | Diagnostic analysis and evaluation of models with 5-40 topics | 4 |
| **Interpretation** | Interpretation of model by describing the topics (stage 1) and creation of broader themes to create the final template (stage 2) | 28 (9 hours per coder) |
| **Total person hours** |  | 40 |

#### Triangulation

We conducted a formal triangulation in order to compare the results from both approaches. Specifically, we performed a methodological and investigator triangulation, as the results from two different analytical approaches performed by two different analysts were compared (Farmer et al., 2006). Two research teams independently analysed the Germ Defence data using the two methods described in the previous sections (MATA and human-only TA). A “convergence coding matrix” (O’Cathain et al., 2010; Tonkin-Crine et al., 2016) was created, and two researchers from these separate teams (LT and PB) independently triangulated the findings from both analyses. The codes were then compared with each other and categorised as either; agreement, complementarity, dissonance, or silence (O’Cathain et al., 2010; Tonkin-Crine et al., 2016). Agreement represented conceptual convergence between the analyses, and complementarity referred to a shared meaning or essence between the findings, but some unique nuances were present. Dissonance represented disagreement between the coding, and silence referred to a finding which was present in only one of the analyses. As such, codes were not considered dissonant with each other when they only represented difference of opinion within the sample, and not between the coding from the two methodologies. For example, the code ‘clear and simple’ from the human analysis was not considered dissonant with ‘wordy and repetitive’ from the MATA because alternative codes were present which agreed, such as ‘information was clear, concise, and easy to understand.’ The two analysts then compared and discussed their decisions and reached consensus on the findings.

## Results

### Person hours

The human qualitative analysis required significantly higher person hours to complete than the MATA (147.5 vs 40). The only stage which less time in the human analysis than the MATA was the final interpretation stage, likely due to the familiarity with the data gained by coding the data ‘by hand’ and the pre-existing coding template. In the MATA approach, the inference of the topics and the classification component of the analysis was conducted by the machine learning model. In this case, the final interpretation phase consisted of the two stages of generating narrative descriptions of the produced topics and following the process of thematic analysis. This was the first time the human coders came into contact with the data and thus this step was the most time-consuming one in the MATA.

### Primary data analysis

The MATA results were centred on what users found helpful and unhelpful about the Germ Defence website. The themes representing what users found helpful were: 1. *Clear and easy to understand*, 2. *Provision of new information and reminders*, 3. *Confirming and Reinforcing*. The themes representing what users found unhelpful were: 1. *Repetitive, simplistic, wordy, patronising*, 2. *Lack of tailoring*, 3. *Various issues relating to usability, content and specific features*. For the human analysis, we found 3 main themes: 1) *layout and language style*, 2) *confidence in how to perform the behaviours*, and 3) *reducing all or nothing thinking*. See online supplementary material 4 and 5 for further detail on the results of the separate primary analyses.

#### Machine-assisted topic analysis process

##### A. What was helpful about the information on the Germ Defence website?

###### Inclusion of the topics in the qualitative analysis

Of 25 topics analysed qualitatively, 22 topics were included in the analysis as they provided substantial insights as expressed by the users’ feedback^1^. See online supplementary material 5 for a ranking of the machine-generated topics in terms of prevalence in the corpus for question A.

##### B. What did you not find helpful about the information on the Germ Defence website?

###### Inclusion of the topics in the qualitative analysis

Of 15 topics analysed qualitatively, 13 topics were included in the analysis as they provided substantial insights as expressed by the users’ feedback^2^. See online supplementary material 5 for a ranking of the machine-generated topics in terms of prevalence in the corpus for question B.

The MATA codes from both corpora were grouped into major themes representing what users found helpful/unhelpful with the Germ Defence intervention (Table 4).

**Table 4:** Summary of the topics (generated by the model, described by human) and the major themes (generated by human)

| MATA themes | MATA codes |
| --- | --- |
| Clear and easy to understand | A1 - Information was clear, concise and easy to understand |
|  | A3 - It was written in simple language making it easy to understand and accessible |
|  | A5 - Useful information that is simple, clear and easy to understand |
|  | A22 - Information was clear, simple, and easy to read and understand |
| Provision of new information and reminder | A2 - Reinforced existing knowledge and practices; helpful extra information and guidance on what more can be done (e.g. at home) that users hadn't thought of before |
|  | A6 - Helpful information users hadn't thought of before; the case studies were helpful |
|  | A8 - Information on how the virus lives and spreads, along with explanation of the link between amount of viral exposure and severity of illness |
|  | A9 - Good reminders and advice on precautions (e.g., social distancing) |
|  | A10 - Helpful information that prompted users to reflect on their current behaviours; scenarios prompted users to provide answers/respond and make plans going forward |
|  | A12 - Helpful new information and advice for in-home mitigation measures; confirmed existing behaviours/measures were right |
|  | A13 - Good reminders and ideas on various mitigation measures, that also confirms existing practices; the option to share the website link with others can be really helpful |
|  | A16 - Good reminders on risks and various mitigation measures |
|  | A20 - Useful, clear and evidence-based information with practical ideas about keeping safe indoors and reducing fomite/surface transmission |
|  | A24 - Provided clarity on length of time the virus lives on surfaces and in air, and highlighted relevant mitigation measures |
|  | A7 - Highlighted the importance of handwashing and reminded users to keep up with handwashing practices |
|  | A21 - Made users think about current precautionary practices and to be more careful |
|  | A14 - Good reminders about hand hygiene, disinfection of surfaces |
| Confirming and Reinforcing | A18 - Helpful information that reinforced existing precautionary practices and shaped attitude towards them, and encouraged further actions to take. |
|  | A25 - Reinforced existing knowledge and behaviour |
|  | A19 - Reinforced existing knowledge/common sense and prompted re-evaluation of behaviours that users have become lax with; highlighted increased risk and importance of reducing large viral exposure |
|  | A17 - Information was helpful and clear while clarifying and confirming what users had understood from health professionals/NHS |
|  | A15 - Confirmed what users already knew |
| Repetitive, simplistic, wordy, patronising | B3 - Wordy and repetitive of what is already well known, while slightly patronizing |
|  | B5 - Visual design slightly under-developed/not user-friendly; information was repetitive or too simplistic. |
|  | B10 - Helpful but repetitive information |
|  | B11 - Did not provide any new information beyond what is already known and is patronizing |
| Lack of tailoring | B12 - Guidance and questions lack consideration for practicalities within families, especially families with young children |
|  | B9 - Unpleasant user experience on the website; Information requires more detail and lack consideration for certain demographics/living situations |
|  | B1 - Some guidance is not practical or sensible based on personal circumstances (i.e., risk and living situation) and latest scientific evidence, and requires harder factual explanation. |
| Various issues relating to usability, content and specific features | B2 - Website not user-friendly (e.g., challenges with navigation) |
|  | B4 - Guidance/questions present too many options but does not consider certain living situations (e.g., living alone) |
|  | B6 - Website not user-friendly as it was difficult to navigate and did not display well on smartphones; some guidance is not realistic/practical (e.g., social distancing at home) as it does not consider its mental health impacts and individual circumstances, while some guidance (e.g., on reducing fomite transmission) is not sensible based on latest scientific evidence. |
|  | B8 - Website not user-friendly as it was difficult to navigate the various options and the web layout made users question credibility of the website; Some information was misleading/confusing (e.g., germs versus virus) while some suggestions are not practical/reasonable (e.g., social distancing within the home) or require more detailed explanations. |
|  | B14 - Rather superficial, lacking explanation and detail for several mitigation measures (e.g., use of masks and disinfectants, hand hygiene); having the option to choose between scenarios was confusing and may not be necessary. |
|  | B7 - Advice to wear a mask at home or socially distance within a home are unreasonable |

Note: The label 'A' refers to codes generated from the question: "What was helpful about the information on the Germ Defence website?" The label 'B' refers to the question: What did you not find helpful about the information on the Germ Defence website?"

### Triangulation

The codes generated from each form of analysis were categorised as either in agreement, or complementary to each other. We found no instances of dissonance or silence within the coding from the two methods. Table 5 presents the full results of the triangulation.

**Table 5.** Results of the triangulation between the human-only analysis and the MATA

| Human-only themes | Human-only codes | Triangulation with MATA codes |  |
| --- | --- | --- | --- |
|  |  | Agreement | Complementary |
| Layout and language style | Clear and simple | A1, A3, A5, A22 |  |
|  | Not enough information | B9, B11, B14 |  |
|  | Not streamlined or sophisticated | B5, B2, B6, B8 |  |
|  | Too repetitive | B3, B5, B10 |  |
|  | Too simplistic/patronising | B3, B5, B11, B14 |  |
| Confidence in how to perform the behaviours | Clear practical advice and troubleshooting is helpful | A2, A6, A9, A10, A12, A13, A20, A24, A7, A21, A14, A18 | B12, B9 |
|  | Feeling informed and reinforced by reliable sources is empowering | A12, A13, A16, A20, A7, A14, A25, A19, A17 | A15 |
|  | Inconsistencies undermine confidence |  | A20, A17 |
| Reducing all or nothing thinking | Trying to perform all the behaviours is exhausting |  | B12, B6 |
|  | Understanding that small changes matter is motivating |  | A8, A21, A19 |
|  | We should act according to risk | B1 | A16, A19 |
|  | Some behaviours are very challenging in certain situations | B12, B1, B4, B6, B8 | B9 |

#### Instances of agreement

There was a high level of agreement between the findings of the human and MATA analyses, particularly for the themes: *layout and language style and confidence in how to perform the behaviours. All of the codes which made up the layout and language style* theme from the human analysis were classified as in agreement with the related codes identified in the MATA. Both methods agreed that Germ Defence users found the website clear to use and easy to understand, but there were a few areas requiring improvement. For example, some users felt that the website did not appear “slick” or sophisticated enough, and that the simple language appeared patronising to some. Some examples of codes classified as in agreement were: ‘clear and simple’ versus ‘information was clear, concise and easy to understand’, and too ‘simplistic/patronising’ versus ‘did not provide any new information beyond what is already known and is patronizing’.

We also found many instances of agreement between the methods for two of the three codes which made up the theme *confidence in how to perform the behaviours* from the human-only analysis. Both methods agreed that many of the participants felt that the website provided important reminders and reinforcement of the recommended behaviours. For example, for those who were already highly adherent to the behaviours, the website provided assurance that they were doing the right thing and encouragement to continue. For those who experienced difficulty performing the behaviours, the website provided practical guidance and ‘real-world’ examples of how the infection control behaviours could be integrated into users’ daily routines. An example of codes classified as in agreement is ‘clear practical advice and troubleshooting is helpful’ from the human-only analysis versus ‘helpful information users hadn’t thought of before; the case studies were helpful’ from the MATA.

Finally, two of the four codes contained within the *reducing all or nothing thinking* theme agreed with codes generated from the MATA. The majority of the agreement here came from finding that some of the behaviours may be more difficult to integrate, particularly for families with young children. Some participants felt that Germ Defence could appear too proscriptive, and placed emphasis on the need to balance the behaviours according to what was deemed practical and necessary for the family to perform to reduce risk. For example, the ‘some behaviours are very challenging in certain situations’ code from the human-only analysis was classified as in agreement with ‘guidance and questions lack consideration for practicalities within families, especially families with young children’ from the MATA.

#### Instances of complementarity

The remaining relationships between the findings of the two methods were judged as complementary and there were no instances of dissonance or silence. Only the theme *reducing all or nothing thinking* contained more codes deemed as complementary than in agreement. Both methods found that users placed emphasis on the need to act according to risk level, and that some of the suggested behaviours could be unrealistic in certain households and/or situations. However, the human analysis placed greater emphasis on the potential mental load of integrating the behaviours, and participants’ interpretations of the viral load messages. The viral load messages encouraged some participants by helping them to understand that even small changes (such as implementing some of the behaviours wherever possible and practical, or that they might tailor their behaviours according to risk) can be effective for reducing their risk of catching COVID and/or illness severity. In contrast, believing that they must perform all behaviours perfectly to avoid virus transmission left some participants feeling defeated. The MATA codes did not wholly reflect these interpretations, and so ‘understanding that small changes matter is motivating’ from the human-only analysis was classed as complementary to codes such as: ‘information on how the virus lives and spreads, along with explanation of the link between amount of viral exposure and severity of illness’ from the MATA.

## Discussion

We aimed to explore the potential value of machine learning analysis techniques to analyse large-scale datasets by conducting a comparison between MATA and traditional thematic codebook analysis using a template approach conducted by humans. We triangulated the results of both forms of analysis in order to highlight the similarities and differences between the two methods, and we compared by the person-hours needed to complete the analyses.

In regard to the primary data, both analyses found that online public health interventions should be clear and concise. For our participants, a slick and professional appearance conveys trustworthiness, and many felt that a website should be uncomplicated and accessible. However, others felt that it seemed overly simplistic and patronising, indicating a need for striking balance when designing interventions targeted to a wide audience. Rather than simply stating the recommended behaviours, our participants highlighted the importance of practical information and real-life examples which aim to help website users envision how the behaviours can be implemented in their own homes. Having the efficacy of the behaviours confirmed by those perceived to be experts empowered participants to act, and reinforced participants’ confidence in their ability to protect themselves and those around them. Finally, our participants indicated that public health interventions should recognise that some of the recommended behaviours can be very challenging in certain situations, and attempting to adhere to all behaviours at all times may not be feasible for many households. Many participants indicated that they would act according to their risk level, and felt that information which appeared overly restrictive and inflexible can leave participants feeling defeated and demotivated. On the other hand, messages which emphasised the concept of viral load helped many participants to understand that making even small changes were worthwhile for reducing viral exposure, and understanding risk reduction as cumulative – rather than absolute – was motivating.

As a result of the triangulation between the two methodologies, we found that the results were very similar, with all codes developed from the MATA classified as in agreement or complementary to the codes developed from the human-only analysis. Where the findings were classified as complementary, this was typically due to slightly differing interpretations or nuance which are likely to be due to the human input to the analyses. For example, the investigator leading the human-only analysis (LT) had analysed previous Germ Defence data, whereas the MATA team had not. It is therefore likely that LT made interpretations based on knowledge gained from previous analyses of Germ Defence data. This particularly seems to be the case for the codes within the reducing all or nothing thinking theme, which were more prominent and developed in the human-only analysis by the Germ Defence team. These concepts were salient to the Germ Defence developers because Germ Defence sought to overcome fatalism about infection transmission. Therefore, some of these differences were likely due to investigator difference, and not methodological difference. That said, the codes from the human-analysis were generally more interpretive than the MATA codes. This is different from the findings from another study which compared human analysis with a different NLP approach. Guetterman et al. (2018) found that while human-only analysis was of higher quality than NLP-only analysis, a combined approach added further conceptual detail and further conclusions than human-only analysis. We did not find this to be the case in the current study, rather, we found that human-only methods yielded similar results to a human-assisted NLP approach.

One potential consideration is that punctuation is removed for the MATA as only words, rather than phrases or sentences, are used as tokens. Due to the purpose of punctuation being to convey and clarify meaning, emphasis, and tone within text, the human coders may have been able to understand nuances within the responses during the early stages of analysis that could have been missed or misattributed by the AI. However, the role that humans play in understanding and interpreting the output of the MATA means that any potential missed meaning should be minimal. Similarly, the topics produced by STM can sometimes be incoherent, or involve multiple seemingly unrelated themes. This would be a major issue if the goal of this method was to conduct an exhaustive and in-depth qualitative analysis of the corpus. However, since the goal of this analysis, and the use case for MATA in general, was to rapidly extract headline insights, this limitation can be mostly overlooked. Nevertheless, researchers should be mindful of these potential issues when they come to interpret the output of the AI.

Due to these considerations, MATA could potentially be seen as a less interpretive method than human-only analysis that is suitable for more descriptive studies of large datasets. Indeed, the concept of information power recommends larger samples for studies with broader, atheoretical, more exploratory aims (Malterud et al., 2016). In order to complete the human-only analysis of a sample of this size, a codebook was created based on previous Germ Defence research, and six research assistants needed to be trained in qualitative analysis. It would not have been feasible to conduct a purely inductive thematic analysis using a large number of coders due to differences in how individuals would interpret and label the data. Other methods of coding large-scale data, such as crowdsourcing though Amazon Mechanical Turk, have been shown to be successful when coding deductively into pre-determined categories (Harris et al., 2015; Hilton & Azzam, 2019; Tosti-Kharas & Conley, 2016). However, in the absence of these categories, such as in more inductive approaches or studies with more exploratory aims, there have previously been few options available to researchers other than to perform human analyses on limited sample sizes. Approaches such as MATA could be a valuable tool for enabling large-scale sampling for these types of studies.

Therefore, MATA offers researchers a less resource intensive and time-consuming approach to conducting broader exploratory studies within large, nationally representative samples. It could be used to augment approaches which tend to adopt more descriptive aims such as codebook TA, coding reliability TA, and content analysis. For analyses such as reflexive TA or interpretative phenomenological analysis (IPA) where researchers wish to engage with the data on a richly interpretive level, and the researchers’ knowledge of the subject matter is considered an important analytic lens, we would not currently consider MATA an appropriate approach based on the current findings.

### Strengths and Limitations

The decision to triangulate human qualitative analysis of Germ Defence data with machine learning analysis was made post hoc, and as such, both teams worked and made analytical decisions independently from each other. Whilst this could be seen as a limitation of the current study, we believe that the high level of agreement and complementarity between the two analyses demonstrate the trustworthiness of using machine learning techniques to analyse large-scale datasets. Despite the independence of the two teams, the MATA was still able to generate findings very similar to the human analysis. As discussed above, machine learning techniques may be best suited to more descriptive qualitative analyses, and so it is likely that the results were consistent due to the descriptive aims of the human analysis and the similarity between the results would likely not have been as great if compared with a more interpretive analysis.

The sample of participants in the current study was largely homogenous. The majority of participants were white, midlife or older, and at higher risk of severe illness from COVID-19. We are therefore unable to draw conclusions from the current study as to the utility of MATA and NLP methodology for the analysis of more diverse, nationally representative samples. Further research is needed to assess how NLP techniques handle more diverse datasets.

## Conclusions

For studies with more descriptive aims, MATA is a trustworthy and potentially valuable tool to assist researchers analyse large-scale open text data. Previously, qualitative approaches have been limited to small sample sizes by its time-consuming nature. By triangulating the results from a traditional human-only thematic analysis with those from MATA, we have shown that both methods generate comparable findings, whilst MATA has the benefit of being less resource and time intensive. MATA could therefore be used to automate the early familiarisation and coding process of more descriptive and less interpretive methods such as codebook analysis or content analysis, especially when the goal is to rapidly extract key topics or concepts from the data for use in a public health emergency. This study contributes to an emerging body of literature into the potential utility of machine learning techniques for use in large-scale qualitative research (Cacheda et al., 2019; Crowston et al., 2012; Guetterman et al., 2018; Leeson et al., 2019; Lennon et al., 2021).

## Data Availability

The datasets generated and/or analysed during the current study are available in the figshare repository, https://doi.org/10.6084/m9.figshare.19514305.

## List of abbreviations

AI: Artificial intelligence
IPA: Interpretative phenomenological analysis
MATA: Machine-assisted topic analysis
NLP: Natural language processing
PBA: Person based approach
STM: Structural topic model
TA: Thematic analysis

## Declarations

### Ethics approval and consent to participate

This study involves human participants and was approved by the University of Southampton Psychology Ethics Committee (ID: 56445). Participants gave informed consent to participate in the study before taking part.

### Consent for publication

All participants consented to the publication of the findings of the current study and anonymised data excerpts.

## Authors’ contributions

LT – design, analysis, writing, triangulation

PB – design, analysis, writing, triangulation

TP – design, analysis, writing

RA – conceptualisation

TC – conceptualisation

BA – conceptualisation, writing

LY – conceptualisation

All authors read and approved the final manuscript.

## Acknowledgements

We would like to thank our voluntary research assistants; Benjamin Gruneberg, Lillian Brady, Georgia Farrance, Lucy Sellors, Kinga Olexa, and Zeena Abdelrazig for their valuable contribution to the coding of the data for the human-only analysis. We would also like to acknowledge Katherine Morton’s contribution to the administration of survey, and James Denison-Day for the construction and maintenance of the Germ Defence website.

## Footnotes

1 The rationale for exclusion of 3 topics from the analysis was: - Topic 4 was deemed incoherent - Topic 11 was described as “Nothing was helpful/Learned nothing new” and hence did not provide a substantial answer to the qualitative question - Topic 23 included mixed issues that were already represented in other themes

2 The rationale for exclusion of 2 topics from the analysis was: Topic 13 was deemed incoherent Topic 15 was described as “Nothing was unhelpful/nothing to dislike” and hence did not provide a substantial answer to the qualitative question

