## Supplementary material 1 for "Applying machine-learning to rapidly analyse large qualitative text datasets to inform the COVID-19 pandemic response: Comparing human and machine-assisted topic analysis techniques"

**Online supplementary material 1**

*A: What was helpful about the information on the Germ Defence website?*

| Topic | %^a^ | Description | Theme | Higher probability words | Higher FREX words | Quotes |
| --- | --- | --- | --- | --- | --- | --- |
| 1 | 2.68% | Information was clear, concise and easy to understand | Clear and easy to understand | understand, just, concis, interest, refresh, better, improv | concis, understand, refresh, just, interest, better, memori | It was clear and concise. Unlike the government's confusing attempts at communication |
|  |  |  |  |  |  | It just made things a little clearer, easy to understand |
|  |  |  |  |  |  | Clear and concise information with no major stumbling blocks |
|  |  |  |  |  |  | The site gave clear, easy to understand information in way that was easy for anybody to understand |
|  |  |  |  |  |  | More informative and easy to understand for all ages |
| 2 | 5.68% | Reinforced existing knowledge and practices; helpful extra information and guidance on what more can be done (eg at home) that users hadn’t thought of before | Provision of new information and reminders | info, thought, someon, done, life, etc, extra | someon, info, extra, done, stop, life, handwash | It clarified and confirmed what I already thought ie when you come into contact with someone, you are also in contact with everyone they've been in contact with etc etc over last 2 weeks. It helped with what more can be done at home |
|  |  |  |  |  |  | Ideas on keeping home safer, cleaning handles etc, I had done that in March but later stopped. I now do it again |
|  |  |  |  |  |  | Extra information in virus life span and possible transmission from parcels delivered etc. Ummmm... |
|  |  |  |  |  |  | Gave valuable extra info on how to how to try and avoid the virus in the home especially if you have someone entering your home |
|  |  |  |  |  |  | Reinforce what I have done for the last three winters to stop getting I'll. ie self isolate, avoid public transport, keep away from those who could pass viruses to me |
| 3 | 6.37% | It was written in simple language making it easy to understand and accessible | Clear and easy to understand | simpl, way, easi, explain, detail, languag, access | explain, simpl, languag, way, quick, detail, logic | The way it was written, context provided, easy language, simple to understand. Logically written. It inspired confidence that the information is a product of academic thought and based in research |
|  |  |  |  |  |  | Very clear (almost unscientific language) makes it accessible to the majority of the population. |
|  |  |  |  |  |  | The way it is worded and done in a way that is simple to read |
|  |  |  |  |  |  | It explains in simple words what we can do to prevent the virus spreading in a way which is accessible to most people ; and also explains virus transmission in simple words too. It is great ! |
|  |  |  |  |  |  | Plain and simple language; Not patronising; Explained nature of the virus and how easy it can be transmitted very clearly |
| 4 | 2.07% | Incoherent | - | contact, bubbl, work, home, covid, famili, time | bubbl, necessarili, contact, issu, rare, mind, work | Nothing, if germs where an issue we would of died out at the beginning. Grems are here part of everything part if us we think we need to eliminate them but all of everything is about harmony and balance. There is a reason that hospitals are the epicentre for MRSA super strains, in elimination of anything balance is lost. The human body is a balance machine harmony |
|  |  |  |  |  |  | The hard facts are rarely publicised. Most of us are intelligent enough to understand but  the Government and BBC cater for the lowest common denominator causing the stupid to be bored and miss important data and the intelligent to switch off, metaphorically and actually |
|  |  |  |  |  |  | I'm afraid nothing that is not already available from the government and NHS, used a little repetitively and polemically, if I'm honest.  Your audience are not necessarily uneducated children: do not talk down to them |
|  |  |  |  |  |  | If I was in an environment where I may come into contact with people who may have the virus or may contract the virus I would find the information very helpful. The Christmas period being one of those situations with extending contact with more family members |
|  |  |  |  |  |  | Useful BUT needs to allow people to personalise their situations...eg I live on my own but my family who visit me are all home working and are all very careful in any contacts they do have....so, in my opinion, have a much reduced risk of catching and passing on Covid . Nowhere in the survey seem to recognise that contact with those who are eg working in shops have a different risk level than those many , many people that are home working ? |
| 5 | 4.28% | Useful information that is simple, clear and easy to understand | Clear and easy to understand | use, easi, clear, set, give, follow, manner | use, easi, set, give, clear, manner, follow | Set it in a clear and easy to understand manner |
|  |  |  |  |  |  | Easy to understand |
|  |  |  |  |  |  | Easy to understand ... clearly set out .. in language I could understand |
|  |  |  |  |  |  | Simple clear sentences |
|  |  |  |  |  |  | It was very clear and easy to follow |
| 6 | 1.97% | Helpful information users hadn’t thought of before; the case studies were helpful | Provision of new information and reminders | case, thought, hadnt, relat, one, studi, laid | studi, case, relat, hadnt, laid, direct, diagram | I hadn't thought about grocery deliveries carrying virus. I will try to leave hard covered items in the cupboard and remove fresh vegetables from packaging before placing in the fridge. I already wash my hands before and after putting food away |
|  |  |  |  |  |  | Finding out that people are most infectious before they develop symptoms. Well laid out options for progressing through the site |
|  |  |  |  |  |  | A clear reminder of why we need to be very careful and conscious of how we behave ourselves and with others. The picture graphs diagrams were good. It is important to tackle the home environment as it is the one hardest to be self conscious about because it is so familiar with preestablished  patterns of behaviour |
|  |  |  |  |  |  | It explained about the amount of exposure being important. It stated clearly the different ways that you can come into contact with a virus. Some of the things we already do at home to protect ourselves were recommended that most people would not consider at home (e.g. 3 day inbox, coupled with decontaminating items that bypass the inbox) |
|  |  |  |  |  |  | Having case studies, and clear expectations of what to do |
| 7 | 3.82% | Highlighted the importance of handwashing and reminded users to keep up with handwashing practices | Provision of new information and reminders | wash, hand, take, import, precaut, everyon, often | wash, hand, forget, precaut, everyon, take, activ | It told me about washing my hands more often  before I do things like eating fruit and snacks which I will be doing from now on |
|  |  |  |  |  |  | Reminding me to wash my hands more than I do already. As time goes on I have not been washing my hands as often as I did during the first lockdown. |
|  |  |  |  |  |  | Giving information about hand washing, 'quaranteeing' items delivered |
|  |  |  |  |  |  | How important it is to wash your hands. |
|  |  |  |  |  |  | Reminded me about when why and how often should wash hands because as time goes on you become more careless |
| 8 | 3.33% | Information on how the virus lives and spreads, along with explanation of the link between amount of viral exposure and severity of illness | Provision of new information and reminders | virus, can, spread, live, long, peopl, surfac | spread, virus, long, prevent, can, tip, live | Previous information given out has said that the virus lives for 24 hours on paper/cardboard (3 days on hard surfaces/plastic)  so any post received has been left for 24 hours, however the information here says the virus can live up to 3 days on paper;  this is new information |
|  |  |  |  |  |  | Information on how viruses spread and how long hey live for outside the body. This helps me to make informed decisions about illness prevention |
|  |  |  |  |  |  | Explaining how long the virus lives and the severity it can have dependent on how much of it you're exposed to |
|  |  |  |  |  |  | Information on how long the virus lives in the air and on surfaces & how it can be prevented |
|  |  |  |  |  |  | Explaining how viruses are spread and how long they can stay on surfaces and in the air. |
| 9 | 5.10% | Good reminders and advice on precautions (e.g., social distancing) | Provision of new information and reminders | good, germ, stay, distanc, social, situat, also | good, stay, germ, social, distanc, term, control | A good run through of the situation and options to safeguard myself and my family |
|  |  |  |  |  |  | General hints and advice, especially about surfaces and the reasons for social distancing |
|  |  |  |  |  |  | Thinking about how I handle deliveries and reminding me of how frequently I should be washing my hands. Simple changes I can make to keep myself safe at home - e.g, socially distancing in my main living room by rearranging furniture |
|  |  |  |  |  |  | Good reminder |
|  |  |  |  |  |  | Good information about how germs are spread and a reminder of the mitigations e.g. clean the home more frequently to reduce the spread of surface germs, and to leave parcels for 3 days before handling. |
| 10 | 6.15% | Helpful information that prompted users to reflect on their current behaviours; scenarios prompted users to provide answers/respond and make plans going forward | Provision of new information and reminders | everyth, make, straightforward, defenc, help, need, info | everyth, fair, defenc, straightforward, make, requir, succinct | Reiteration of the rules I follow & those I've been slacking.  Overall I realise I need to be more vigilant and proactive in ensuring I play my part in helping to combat contracting & passing on germs of any sort |
|  |  |  |  |  |  | Highlighted in home defences that have not had a high profile compared to workplace or retail. Good story telling and an attempt to make it real and relevant |
|  |  |  |  |  |  | Explanation of "why" you would take the measures ; And fairly straightforward to navigate. Succinct and graphic enough; Summary a useful addition for passing round |
|  |  |  |  |  |  | It's made me and my family review our behaviour regarding the virus and go back to the measures we took at the beginning of lockdown in March. |
|  |  |  |  |  |  | Helpful as a round-up of, and addition to, the information that has dribbled through the government and media over the last year |
| 11 | 2.84% | Nothing was helpful/Learned nothing new | - | noth, new, realli, need, educ, just, anyway | noth, educ, anyway, new, realli, touch, constant | Nothing I don't agree with what is written as u are now saying to distance from our family and I ain't going to not hug my son that lives with me or my partner |
|  |  |  |  |  |  | Nothing. I watch the news, am a key worker and I have researched. I have learnt nothing new from this. |
|  |  |  |  |  |  | At this stage I didn't learn anything new. However I do follow research articles. Nothing |
|  |  |  |  |  |  | n/a - there was nothing new that isn't constantly being said in the media. Just more reinforcement. |
|  |  |  |  |  |  | Nothing really. Sorry |
| 12 | 4.30% | Helpful new information and advice for in-home mitigation measures; confirmed existing behaviours/measures were right | Provision of new information and reminders | keep, safe, open, window, day, hous, deliveri | open, window, keep, safe, straight, deliveri, hous | Never wore a face mask in the house but will when receiving a delivery |
|  |  |  |  |  |  | The information about how to use, handle & treat masks, & how to keep surfaces & door knobs clean, & what do do about receiving deliveries. I already knew this information & have been teying to carry it out, but it was good to read it again, & to be able to read it out to my husband so that he knew it was coming from a reliable source, as he's not as diligent as i am & tended to get angry when he saw me doing it or if i said we need to do these things to keep ourselves safe.  I would have liked more information on what to do if another member of the family wants to visit |
|  |  |  |  |  |  | Remembering to store parcels before opening them |
|  |  |  |  |  |  | Information and advice that I'd not heard before: that coronavirus lives externally for three days, that wearing glasses helps and leaving alone or cleaning mail deliveries |
|  |  |  |  |  |  | The advice about keeping windows open, because this has received little publicity in a home context |
| 13 | 3.46% | Good reminders and ideas on various mitigation measures, that also confirms existing practices; the option to share the website link with others can be really helpful | Provision of new information and reminders | gave, protect, idea, other, lot, basic, re | gave, basic, lot, other, idea, protect, websit | It verified that for the last 9 months is correct and it's something I been taught as a child re cleanliness |
|  |  |  |  |  |  | Re iterate what one needs to do to protect themselves and family. |
|  |  |  |  |  |  | How it transmits.  How to prepare when seeing others. The benefit of not having others in your personal space |
|  |  |  |  |  |  | Sitting opposite even 2 metres apart not a good idea, I thought we would be ok as one person not been to a shop etc for months, another every 9 days, 3rd paper shop most days. Good reminder re washing hands on returning home 2/3 always do so. Will antibacterial internal door handles more often and kitchen work surfaces as usually only wash down once a day and damp clean as used |
|  |  |  |  |  |  | The examples of how some people are dealing with disease prevention at home gave some useful ideas. The option to share the link with family and friends is a good idea too. |
| 14 | 4.69% | Good reminders about hand hygiene, disinfection of surfaces | Provision of new information and reminders | remind, thing, clean, becom, disinfect, surfac, etc | remind, thing, becom, clean, disinfect, alon, correct | the reminder that soap and water was more effective than water alone or gel |
|  |  |  |  |  |  | reminding about cleaning surfaces, buttons, doorknobs etc. |
|  |  |  |  |  |  | Reminder about opening windows for a while, having the visitors in that room only. Leaving parcels and letters alone for a few days, but if urgent, to clean over parcel or letter with a cloth and disinfectant a good idea. Reminder to clean as you go in the house |
|  |  |  |  |  |  | Reminders about cleaning surfaces and parcels etc |
|  |  |  |  |  |  | It reminded me that I should also wash my hands after I've blown my nose |
| 15 | 5.32% | Confirmed what users already knew | Confirming and reinforcing | alreadi, know, knew, confirm, reiter, yes, told | knew, alreadi, know, confirm, most, prompt, reiter | It mostly confirmed what we knew already. It was good to see the 72-hour period for quarantining deliveries: it's what we do already (with the exception of food that has to go in the fridge). |
|  |  |  |  |  |  | Reiterating what I already knew. |
|  |  |  |  |  |  | It confirmed what I already knew. |
|  |  |  |  |  |  | Mostly confirmed what I know. |
|  |  |  |  |  |  | It reaffirmed what I already knew |
| 16 | 2.89% | Good reminders on risks and various mitigation measures | Provision of new information and reminders | advic, awar, practic, risk, given, remind, stori | awar, advic, given, practic, stori, risk, mix | Reminded me of the protocol and highlighted a few other areas that we can hopefully put into practice over the bonkers Christmas 5 days |
|  |  |  |  |  |  | Raising awareness of practical measures to adopt and a reminder of day to day risks. |
|  |  |  |  |  |  | Reminded me of the risks I face daily and perhaps I catched up with some advices I knew but I did not implement quite often |
|  |  |  |  |  |  | The examples of what people did in their homes (adam's story for example) rather than the list of information which is stuff that I've seen before. Helpful to be reminded that the accumulation of contacts with the virus means that you're likely to be affected more |
|  |  |  |  |  |  | Timely reminder of some actions I was following. Good advice on some areas I had previously ignored - eg packages arriving |
| 17 | 4.47% | Information was helpful and clear while clarifying and confirming what users had understood from health professionals/NHS | Confirming and reinforcing | help, explan, peopl, found, avoid, health, state | explan, help, found, though, health, profession, state | Even though I am already aware of the information, I still found it helpful. I think the information about leaving deliveries outside for a few days was helpful, as many people do not know it (or follow it). However, I am a bit confused about whether 3 days is enough, as some research says that the virus survives for a much longer time on plastic and metal |
|  |  |  |  |  |  | It was clear. It clarified what I had understood from health professionals. It helped make me feel secure in choices I had made, whilst woolly thinkers had been trying to persuade me that I was being too careful. In fact, I was not being quite careful enough. |
|  |  |  |  |  |  | Clearly stating what should and shouldn't be done. I'd forgotten about the leaving of parcels for 72hours |
|  |  |  |  |  |  | It stengthened the advice I have been sent by NHS.  I am in the exteme category. |
|  |  |  |  |  |  | The information was described in clear ways with no unnecessary jargon and without being too overwhelmingly wordy |
| 18 | 4.37% | Helpful information that reinforced existing precautionary practices and shaped attitude towards them, and encouraged further actions to take. | Confirming and reinforcing | action, measur, suggest, see, best, differ, take | action, point, best, suggest, measur, see, right | It reinforced my belief that the higher intensity of the viruses I am in contact with, the more chance I have of contracting the disease and the worse my illness might be.\nIt also reinforced my determination to continue the measures we already use to decontaminate as much as possible that we bring into our home including our hands, and to consider improving on our practice of these measures. |
|  |  |  |  |  |  | Brings home the importance in a none judgemental way whilst encouraging the right habits |
|  |  |  |  |  |  | Made you consider precautions that you have started to take for granted |
|  |  |  |  |  |  | I identified what I dont do right and what I should do |
|  |  |  |  |  |  | Confirmation that my present regime is on the right track |
| 19 | 6.80% | Reinforced existing knowledge/common sense and prompted re-evaluation of behaviours that users have become lax with; highlighted increased risk and importance of reducing large viral exposure | Confirming and reinforcing | read, common, sens, re, reduc, exampl, risk | sens, common, read, enforc, larg, re, view | I thought I knew it all but it made me re evaluate my knowledge and I got abit relaxed over my germ control as my quiz answers showed so I will step it up again after reading this |
|  |  |  |  |  |  | Reinforces the key messages |
|  |  |  |  |  |  | It reminded me of some things I did at the start of the pandemic, which I have become less rigorous about, for example, leaving some things that are delivered to reduce the risks of virus transmission |
|  |  |  |  |  |  | Yes windows open whilst common sense it re enforced the thought |
|  |  |  |  |  |  | The information was largely practical and easily understood. I felt the examples of people being careful made it seem achievable. The clear statement that reducing the amount of virus you might be exposed to was well made. This seemed possible to do, although the thought of disinfecting all hand contact surfaces in the house even daily and any food bought is more than I have been doing and I feel I have been very careful since the start of the pandemic. Good to put forward that this is training for life and would help reduce a range of viral infections |
| 20 | 5.16% | Useful, clear and evidence-based information with practical ideas about keeping safe indoors and reducing fomite/surface transmission | Provision of new information and reminders | mask, well, wear, indoor, home, famili, use | indoor, mask, wear, non, well, scientif, testimoni | Digestible info, ability to share easily on social media, I learned things even as a Chemistry graduate I didn't know. Liked the info about creating safe places even in non-symptomatic homes, to protect me/rest of household |
|  |  |  |  |  |  | It gave me new ideas about how to isolate if we were ill and the evidence to back up why. I had Covid-19 in April and my husband refused to be in a separate room, he didn't catch it luckily but we did all isolate from the outside. |
|  |  |  |  |  |  | Information on if a member family had symptoms but no one else. How to isolate them and to try and protect rest of family |
|  |  |  |  |  |  | The need to wear a mask even indoors if family visit. Need to have windows open. I hadn't realised that the longer you are in the same space as a person with Covid the more of the virus you absorb |
|  |  |  |  |  |  | Very useful. It was a gentle reminder, after being isolated for so long. I have always opened the postal delivery as soon as it arrives. It is a vital link to the outside world |
| 21 | 3.38% | Made users think about current precautionary practices and to be more careful | Provision of new information and reminders | think, made, home, care, load, viral, peopl | made, think, load, care, home, viral, relev | Reinforced idea that viral load was relevant in the severity of disease if contracted, and that handwashing was key to reducing viral load. I had not realised this virus was more infectious BEFORE patient becomes symptomatic - I will be even more careful now! |
|  |  |  |  |  |  | It made me think about the things I'm doing at home and away right now so |
|  |  |  |  |  |  | It made me think about being more careful in my home |
|  |  |  |  |  |  | I thought I was doing pretty well with my levels of germ defence in the home but it made me realise there were areas I could do still do better with |
|  |  |  |  |  |  | It brings back into focus good practice that was more common during the first National Lockdown that may have been forgotten or seen to be less pertinent now |
| 22 | 3.51% | Information was clear, simple, and easy to read and understand | Clear and easy to understand | inform, clear, clariti, general, provid, easi, simplic | inform, clariti, clear, simplic, illustr, provid, general | Easy to read information |
|  |  |  |  |  |  | Giving clear information |
|  |  |  |  |  |  | It was plain English and no page was overloaded with information |
|  |  |  |  |  |  | I found the information to be clear, easy to understand and perfectly sensible. Much of it is information that is already known (and applied) by the majority of sensible and responsible people |
|  |  |  |  |  |  | Information is very plaintext and clear to understand |
| 23 | 3.79% | Mixed issues that were already represented in other themes | - | much, didnt, need, littl, might, realli, thought | didnt, much, might, elder, ocd, need, littl | Not much really. My wife and I are retired and since the lock down we haven't been going out much and always stay 2 Metres from others |
|  |  |  |  |  |  | The information is straightforward and can easily be adopted as part of everyday life sit doesn't need anything more than common sense and the PPE that you'd use daily |
|  |  |  |  |  |  | Accessible and I feel my elderly relative may read it and take it onboard because she doesn't always take onboard what I say. |
|  |  |  |  |  |  | There were many facts that I didn't know, despite tuning in daily to information media. |
|  |  |  |  |  |  | It reinforced the information that was already available and really for me did not go beyond what has already been recommended. I was in Spain in January and realised the implications straight away. My wife says I am boring as I have been telling people about this for years. I am a firm advocate of green matters |
| 24 | 2.38% | Provided clarity on length of time the virus lives on surfaces and in air, and highlighted relevant mitigation measures | Provision of new information and reminders | time, can, surfac, length, aspect, covid, current | length, aspect, time, current, remain, asid, routin | It informed me about some aspects that I have not seen or heard elsewhere, such as the length of time the virus can remain alive on surfaces. |
|  |  |  |  |  |  | clarity about some issues eg about length of time for virus to remain alive on surfaces and in air |
|  |  |  |  |  |  | The length of time that the virus can last on different surfaces |
|  |  |  |  |  |  | Most things, but in particular the time that the virus could reside on hard surfaces |
|  |  |  |  |  |  | It was very comprehensive about the covid virus and how easily it can be spread.  It highlighted things I don't do very well - such as, although I do clean my mobile phone, I almost certainly don't do it sufficiently often.   It is also too easy to find oneself getting closer to people without realising it and when the restrictions eased somewhat it was difficult with Grandchildren to always maintain that distance. |
| 25 | 1.17% | Reinforced existing knowledge and behaviour | Confirming and reinforcing | reinforc, knowledg, infect, protect, use, month, idea | reinforc, knowledg, infect, month, ago, either, protect | Reinforced existing knowledge and behaviour |
|  |  |  |  |  |  | It reinforced all the ideas of protection that I already had but it was a useful tool to persuade my husband to be more careful as he tends to find my reminders irritating. He thinks me over zealous |
|  |  |  |  |  |  | It reinforced our knowledge |
|  |  |  |  |  |  | Reinforced and confirmed much of the information I had already knew about coved-19 and how to protect oneself from being infected |
|  |  |  |  |  |  | It reinforced  the knowledge  I already had |

^a Topic prevalence in the corpus as percentage^

*B: What did you not find helpful about the information on the Germ Defence website?*

| Topic | %^a^ | Description | Theme | Higher probability words | Higher FREX words | Quotes |
| --- | --- | --- | --- | --- | --- | --- |
| 1 | 7.94% | Some guidance is not practical or sensible based on personal circumstances (i.e., risk and living situation) and latest scientific evidence, and requires harder factual explanation. | Lack of tailoring | n, one, risk, answer, home, question, person | n, answer, risk, one, hard, question, realist | I wanted to say I understand the info about keeping apart in the home however we don't all have the space. We can't keep 2 metres apart as our family contains 3 adults in a small terrace with a poky living room plus we're all having to work/study from home. |
|  |  |  |  |  |  | Psychologically it is hard to imagine wearing a mask at home when no one has symptoms.  Equally it could be said to be sensible. Esp now we are approaching winter.  However I am not sure how convinced people would be to create these barriers Ami get family members, not least the person they share a bed with |
|  |  |  |  |  |  | I wasn't sure the information was up to date or helpful in terms of personal risk. Leaving cardboard for 3 days? Wearing a mask when you have no risk factors, work from home, don't go out and live with one other person doing the same? |
|  |  |  |  |  |  | The information is fundamentally wrong. It doesn't reflect the real or natural world or how humans live and thrive |
|  |  |  |  |  |  | Could have had something about balancing working from home with partner who goes out to work? |
| 2 | 4.10% | Website not user-friendly (e.g., challenges with navigation) | Various issues relating to usability, content and specific features | page, site, navig, inform, option, click, back | navig, page, applic, click, site, text, start | Unnerving having different options to select. If I click on this button it takes me somewhere, but what about the other buttons I didn't click? What have I missed? Potential anxiety. |
|  |  |  |  |  |  | Navigation is a bit 'clunky' requiring selection of options, it's not very easy to determine what you haven't seen on the website |
|  |  |  |  |  |  | Going to a different site was bit confusing. Said something like 4 pages, then clicked onto connected site and several further pages. Wd be better if simply condensed into one article, perhaps with optional links for further info listed at the end |
|  |  |  |  |  |  | When I started the website I was given multiple options on one page. Having selected one option I was led through various pages of information and questions without being returned to the starting page when finished. I do not know if by taking the route that I did I covered all of the material on the website or not. |
|  |  |  |  |  |  | ....the big BUT for me is the website design, layout and navigation  The presentation is - naturally - from an academic environment but the website has to compete for our attention in a commercial - sometimes anti C19 caution- environment(s).  Take this test. Put a typical GD page up on your screen. What do you view? A mass of text ...Yes? No room for the mind and eye to breathe to help digest the information. Needs more graphics/pictorial content and information ( words ) reduced to shorter key statements.  What is the point of the 'Return to Start' and 'Home page' navigation? The terms are too similar. The navigation is trying to get us to read key statements before the subdivision between the ill and handwashing page split.   This is not entirely successful, to my mind, as, and if, you hit the Home button you are then taken back to the key information headings - two of which you might have read!  Start with the home page links alongside key bold statements from the above intro section.   You have used a free source for graphics but they appear slightly childish in appearance and do lend the site authority.   ( I appreciate there might be a very small budget for graphics ) |
| 3 | 9.58% | Wordy and repetitive of what is already well known, while slightly patronizing | Repetitive, simplistic, wordy, patronising | bit, alreadi, found, seem, littl, everyth, patronis | bit, awar, patronis, wordi, knew, found, everyth | Too wordy, the chatty style took too long to get to the point and I found this slightly patronising |
|  |  |  |  |  |  | It was repetitive. Much of the information has already been well publicised. I found the'plan' rather patronising |
|  |  |  |  |  |  | It took too long to read and seemed a little repetitive |
|  |  |  |  |  |  | That there was nothing new that isn't constantly being said in the media. Just more reinforcement |
|  |  |  |  |  |  | To start with it seemed a bit obvious  but as I say above, it soon became clear that I could learn and need to improve my hand cleaning.. maybe you could say something like, maybe you’ll be surprised! |
| 4 | 5.02% | Guidance/questions present too many options but does not consider certain living situations (e.g., living alone) | Various issues relating to usability, content and specific features | good, time, need, option, live, know, far | good, close, elder, far, need, time, option | I was unsure whether those living alone (a large percentage of the population) were fully identified |
|  |  |  |  |  |  | The suggestion that people wear glasses when for example shopping, as the eyes are points of entry for coronavirus. It doesn't work for me. I can hardly see because of the fogging I get while wearing my mask. Last time water streamed down my glasses, presumably from my breath.  "Do you live on your own?", doesn't really cover my situation as I am a part-time carer for an elderly relative who lives nearby, including giving close care at times. Also I am in the bubble of a friend and his mother where she is very elderly and is bed bound with alzhiemers.  Lack of environmental consideration and contradictions. Is it really necessary to throw plastic bags that masks have sat in away, given that the site also claims that coronavirus does not exist on surfaces for longer than 72 hours? Why can't plastic bags be washed in hot water, left to dry and quarantined? |
|  |  |  |  |  |  | Too many 'boxed 'options |
|  |  |  |  |  |  | Too fussy with all the options and alternatives. A simple "must read" list would be more straightforward |
|  |  |  |  |  |  | My computer did not download the full extent of the options and whilst I did think there was another option to the far right of the table, I was only able to read it in print mode! |
| 5 | 5.59% | Visual design slightly under-developed/not user-friendly; information was repetitive or too simplistic. | Repetitive, simplistic, wordy, patronising | much, read, lot, make, quit, peopl, inform | much, quit, lot, simplist, read, phone, pictur | A great deal of the information was repeated ad nauseam, in my opinion making the process take longer than necessary |
|  |  |  |  |  |  | It was a bit too simplistic, quite a lot of it was obvious standard advice, such as cleanliness to avoid contamination and spread of germs |
|  |  |  |  |  |  | Too much to read. Information could be more concise |
|  |  |  |  |  |  | I found the website practical, useable and informative. However, I found the visual design a bit under-developed - it seemed to not be a priority. I couldn't help feeling that a further evolution of the visual design of the site might extend the reach of the project |
|  |  |  |  |  |  | The picture with lots of people showing the spread It was a complicated picture. It could have been better |
| 6 | 2.33% | Website not user-friendly as it was difficult to navigate and did not display well on smartphones; some guidance is not realistic/practical (e.g., social distancing at home) as it does not consider its mental health impacts and individual circumstances, while some guidance (e.g., on reducing fomite transmission) is not sensible based on latest scientific evidence. | Various issues relating to usability, content and specific features | health, necessari, didnt, time, back, can, part | health, necessari, mental, part, coupl, couldnt, didnt | Your information on fomite transmission of the virus may not be accurate. A recent Australian study suggested that this RNA virus can live on objects outside the body for up to 28 days. Virus living 72 hrs is controversial. Also recent research in the US suggests that the when coughing the virus can travel 6 ft not 2 ft. There is also controversy over the type and safety of the mask worn for protection of self and others. |
|  |  |  |  |  |  | The style of the website was very homespun. Did not display well on a phone. It is important that the presentation/visuals are smart and consistent. |
|  |  |  |  |  |  | Didn't find the way the information was set up. Also felt focus on cleaning/quarantining parcels etc was excessive considering limited evidence of benefits and is more likely to cause anxiety than serve any useful purpose |
|  |  |  |  |  |  | Having gone through it once, it was not easy to get back to some parts I wanted to read a second time |
|  |  |  |  |  |  | I felt that some of the "go to" buttons were confusing and added a layer of complexity that was not necessary.  I am a little unsure about the suggestion that seems to be implied that face masks should be worn at all times when in the same room as someone else in your home , including people you live with and your support bubble - I don't feel comfortable with this and I would also be concerned about health consequences of wearing a face mask for prolonged periods - |
| 7 | 7.69% | Advice to wear a mask at home or socially distance within a home are unreasonable | Various issues relating to usability, content and specific features | home, advic, wear, mask, isol, self, husband | isol, na, self, wear, home, husband, advic | It's not reasonable to expect you to social distance from your children or to wear a mask in your home |
|  |  |  |  |  |  | Difficult to self isolate from husband in same home. |
|  |  |  |  |  |  | The wearing of face masks inside the home |
|  |  |  |  |  |  | Staying separate from my wife in the house would be intolerable |
|  |  |  |  |  |  | The draconian measures suggested. The story about the husband who isolated inside his own home was very depressing. If the child had cancer or similar it might have been wise, but this wasn't clear. If Covid is that dangerous he shouldn't have come home at all. But it isn't! |
| 8 | 2.12% | Website not user-friendly as it was difficult to navigate the various options and the web layout made users question credibility of the website; Some information was misleading/confusing (e.g., germs versus virus) while some suggestions are not practical/reasonable (e.g., social distancing within the home) or require more detailed explanations. | Various issues relating to usability, content and specific features | virus, go, say, like, simpl, number, read | number, particl, im, simpl, critic, say, sever | I didn't like having to pick one option out of the four which best applied to me I would have liked to have picked the most relevant option, be taken through that info and then back to all 4 options to read the others if I wanted to, as I'm sure there are some usual points in all sections.  Also I realise that the website was probably designed to be accessible which is important but when I was first brought to the website my first instinct was that clicking to read the info would give me a computer virus and I almost didn't proceed. I went back to check that the link definitely came from my doctor's surgery. This is mostly because the website really didn't feel at all official and felt very dated and basic. I realise a simple website doesn't necessarily mean that it's nefarious but it felt like the kind of site someone could create on their own and just put 'University of Southampton' on |
|  |  |  |  |  |  | All the things you suggest doing in the home.  Am I supposed to wear a mask when I go to sleep at night as I sleep in the same bed as my husband - less than 2 metres away?  Or are you suggesting separate bedrooms whilst this virus is still in the country?  Frankly, I think the advice was laughable.  I'm not keeping items that come in the post into quarantine for 3 days either as there's no proof the virus can live that long on the items.  I stick to normal household cleaning and personal washing which I'm sure is adequate |
|  |  |  |  |  |  | Page 2 was confusing. Were you referring to  just one virus’ as being a virion or one type of virus such as influenza of coronavirus etc. I understood what you meant but I had to read it a couple of times to make sure.  Telling people to use a disinfectant without explaining that some disinfectants do not kill viruses is not helpful. I would like to see the general term, germ, defined with an explanation that  bacteria and viruses which are under the umbrella term germs, are not the same. So to disinfect correctly the correct type of product has to be selected otherwise it is useless |
|  |  |  |  |  |  | A little confusing number of additional choices for extra info |
|  |  |  |  |  |  | Misinformation about the potential severity of symptoms being related to the number of virus particles (NB not using the term "viruses" - which implies number of virus types) an individual is exposed to.  There is no reputable evidence that the number of particles, beyond critical infection potential, has any effect on the severity of an infection, and it is misleading to suggest so.  Once it has gained entry to the body cells it needs in order to replicate, the number of virus particles in a sufferer can increase enormously, so that the initial number of infectious particles becomes almost irrelevant.  It is incorrect to suggest that people can protect themselves from more severe disease by reducing their exposure, implying that if they are only exposed to a small number of virus particles, they are likely to have only mild symptoms of disease.  Only by avoiding critical infection potential can they avoid whatever course the disease may take in themselves, personally - which is likely to be a function of their biology, not the virus's biology.   Critical infection potential is at the moment unknown, but may be as few as 50 particles, depending on an individual's first line immunological response |
| 9 | 5.13% | Unpleasant user experience on the website; Information requires more detail and lack consideration for certain demographics/living situations | Lack of tailoring | peopl, hand, understand, wash, need, might, basic | understand, fit, basic, wash, age, hand, peopl | You have not thought through the categories of people sharing lives at home without support or third party shielding considerations |
|  |  |  |  |  |  | People have many different circumstances. We are both retired and at home and keeping distant from other people, but not each other. This would be different if we had more people in our environment |
|  |  |  |  |  |  | The website assumes that we all live with someone. We don't! There should have been a section for the millions of us who live alone, especially for older people who are not very able with websites, or can't access them, and who like me are permanently shielding from Covid-19 because they are so vulnerable due to other health problems. Older people are more fearful than younger people about  going out even when it is permitted. More reference to age in the construction of the survey would help, especially in the beginning of it. |
|  |  |  |  |  |  | Someone urgent needs to help people alone with dementia understand - my 82 yr old mother in law has NO understanding despite daily face time calls all sees is the new relating to other people - how often do you see a direct reference to her age group to get help |
|  |  |  |  |  |  | I wish there was more detail about how to protect from the virus e.g. Masks - wash after each use, don't touch the front, take off from the ear straps, ensure nose and mouth covered, bend the top of the mask to fit over the nose if there is a strip; can disposable masks be used again after so many days Hand Gel - should be at least 60% alcohol and rubbed in hands as you would wash your hands Space - is it correct that you will not catch the virus unless you are within two metres of someone for more than 15 minutes inside or outside Shopping - is it necessary to wash/wipe down everything and, if so, will ordinary wet wipes kill the virus? Clothing - do I need to change my clothes if I have sat on a seat used by others, e.g. waiting rooms? |
| 10 | 8.10% | Helpful but repetitive information | Repetitive, simplistic, wordy, patronising | help, inform, someth, thought, seem, link, level | help, someth, often, inform, approach, link, level | That it is all repeated information that we have been told since March 2020 |
|  |  |  |  |  |  | Some suggestions a bit unrealistic/patronising, so will not be forwarding the link to daughters in law. An example would be implication that hands should be washed every time you sneeze or cough - as someone who suffers from a frequent runny nose (often triggered by eating or drinking), and an intermittent cough (often initiated by dry, crumbly food, or sometimes an air temperature/humidity change), I would be washing my hands almost continuously! Another would be the wording around dealing with children, for example with social distancing. |
|  |  |  |  |  |  | It seems likely that the approach would have been helpful in March 2020, and is now nearly at the end of its helpful life : as if the people following GermDefence are being superior to those who have yet to adopt it. The pages are too full of information. Some pages promise information which is then not given but replaced with new information : for example, something about four approaches which were not then clearly enumerated or illustrated. They hid behind other headings, with buttons to press. Repetitive information on other pages led me to give up. |
|  |  |  |  |  |  | Repeating the same information |
|  |  |  |  |  |  | In my case I had this knowledge;   But for others I think it would all be helpful |
| 11 | 6.01% | Did not provide any new information beyond what is already known and is patronizing | Repetitive, simplistic, wordy, patronising | t, thing, anyth, alreadi, think, don, see | t, didn, anyth, don, tell, thing, survey | It didn’t tell me anything I don’t already know. The idea of keeping apart from my own household is ridiculous, it’s bad enough that I can’t see anyone else. |
|  |  |  |  |  |  | Didn’t tell me anything by I don’t already know |
|  |  |  |  |  |  | The website is extremely patronising and assumes that we are all stupid. It seems that this was aimed at Primary School children and not adults who have had at least a standard Secondary School education. |
|  |  |  |  |  |  | Long winded way of saying keep clean, don’t go anywhere, and keep people away |
|  |  |  |  |  |  | I felt anxious reading scientific recommendations and knowing I won't be able to implement most of it in my home. There's a real lack of practical advice for parents. No mention of wiping toys regularly, avoiding play dates or out of school clubs, removing/washing school uniform as soon as they come home, but does tell you live in separate rooms where possible! Advising parents to build a den and play i-spy across a room for a two week stretch of isolation is frankly ridiculous. Oh and patronizing - please remember not to open windows where there's a danger of kids climbing out! Or 'it's a good idea to leave disinfectant/cleaning things near where they'll be used -  but not if you have children around! It's impossible to put most of the suggested measures in place unless you are a 2-4 adult household. Would be better to shut the schools to reduce social contact with germ ridden peers than live socially distanced inside the home - at least kids would have a sense of 'normal' at home and not have to have a bed time story with a masked/goggled parent. How do you spoon feed a toddler or brush teeth from 6 ft away? |
| 12 | 4.00% | Guidance and questions lack consideration for practicalities within families, especially families with young children | Lack of tailoring | famili, household, work, common, symptom, sens, school | common, famili, safe, young, symptom, household, sens | I don’t think the questionnaire or plan’ considered families from a practical perspective. We have a young family so unless either myself or my partner is diagnosed with coronavirus or we have a fever symptom we would not social distance from each other in the home. If my child had a symptom we would also not socially distance. I don’t think the questionnaire accounted for the impact the mitigations could have on Children if the parent followed all of the mitigations suggested. It could also cause someone stress if the felt they had to go against guidance in order to look after their family. Perhaps a questionnaire/plan more specific to families with young children could be added as an option |
|  |  |  |  |  |  | It does not clearly address the complexity of the home where a family group includes a carer who is part of the family. For example how can you socially distance when you are cooking and providing a meal for a member of your family or when you have to provide an act of care |
|  |  |  |  |  |  | Unrealistic to expect families to follow guidance unless someone is showing symptoms |
|  |  |  |  |  |  | It was difficult to answer the questions precisely as the 'family' in the survey was not fully reflective of our family setup |
|  |  |  |  |  |  | No relation to having babies or young children especially if breastfeeding |
| 13 | 4.27% | Incoherent | - | none, relev, virus, two, other, thought, make | none, relev, excel, two, thought, minimis, cough | First two examples were men, put me off till I scrolled and found the bottom two were women. |
|  |  |  |  |  |  | The explanation of  viruses’. I assume you meant viral load. |
|  |  |  |  |  |  | Examples. Not necessary |
|  |  |  |  |  |  | I think we are missing areas of contact-  Ie.  Touching and using drinking vessels- rims of cups and glasses and then touching clean ones-  I feel this is VERY relevant in hospitality sector and observed this being done! |
|  |  |  |  |  |  | It failed to make any mention about  a. Living a healthy or healthier lifestyle, minimising unhealthy habits like smoking and drinking, and taking regular (daily) exercise, if only to inhale plenty of fresh air.  b. Consuming a balanced diet, in particular avoiding foods that subsequently make your body feel heavy, even fatigued and bloated, however one may enjoy their taste. c. Minimising the intake of patent (non-prescribed) medications that may have unexpected side affects, for example hot lemon juice sweetened with honey is much healthier and more effective that Lemsip et all, while I had to wean my mother off strong cough linctus to which she had become addicted. d. If you are unable to rid yourself of a harsh cough from a flu or viral infection, seek from your GP a prescribe, antibiotic cough throat rinse (Difflam). A persistent cough can substantially degrade your condition, especially if it deprives one of proper sleep. |
| 14 | 4.85% | Rather superficial, lacking explanation and detail for several mitigation measures (e.g., use of masks and disinfectants, hand hygiene); having the option to choose between scenarios was confusing and may not be necessary. | Various issues relating to usability, content and specific features | use, differ, confus, think, alway, section, rather | scenario, alway, confus, differ, rather, use, disinfect | I felt it was rather superficial. For example it talked about "using disinfectant to clean taps, etc, but gave no advice on what disinfectants are suitable. Many are pretty useless in removing viruses I believe! |
|  |  |  |  |  |  | Confusing as to which scenario to select. |
|  |  |  |  |  |  | Having to pick the different scenarios when they have the same information, such as how to protect other members of the household if you think you might have coronavirus vs if someone else thinks they have it. The separation is useful if the scenario actually applies but if you are just looking for general information the choice is not needed |
|  |  |  |  |  |  | Disinfectant was mentioned but I think I've always seen alcohol or bleach.  And I'm not sure that all disinfectants work.  I used bleach or Milton that you dilute and must be refreshed every day.  I was a little confused whether it meant lots of Covid 19 virus or different types viruses. I found the different choices confusing.  I'm not sure if they could have been displayed in a different fashion.  Possibly show what the choices were and then have the option to choose.  I don't think you automatically went on to be able to access the next type of scenario automatically |
|  |  |  |  |  |  | I had to retrace through sections to find the section on masks   I have made my own washable mask with liner.  Based on a hospital type that does have ear loops.  It could be useful to some people |
| 15 | 23.29% | Nothing was unhelpful/nothing to dislike | - | noth, realli, unhelp, box, fine, stuff, mind | noth, unhelp, fine, box, negat, stuff, iphon | Nothing springs to mind |
|  |  |  |  |  |  | Nothing was unhelpful |
|  |  |  |  |  |  | There was nothing on the website I disagreed  with |
|  |  |  |  |  |  | Nothing really |
|  |  |  |  |  |  | Nothing was unhelpful |

^a Topic prevalence in the corpus as percentage^

Figure 1. Mind map of the MATA themes and topics

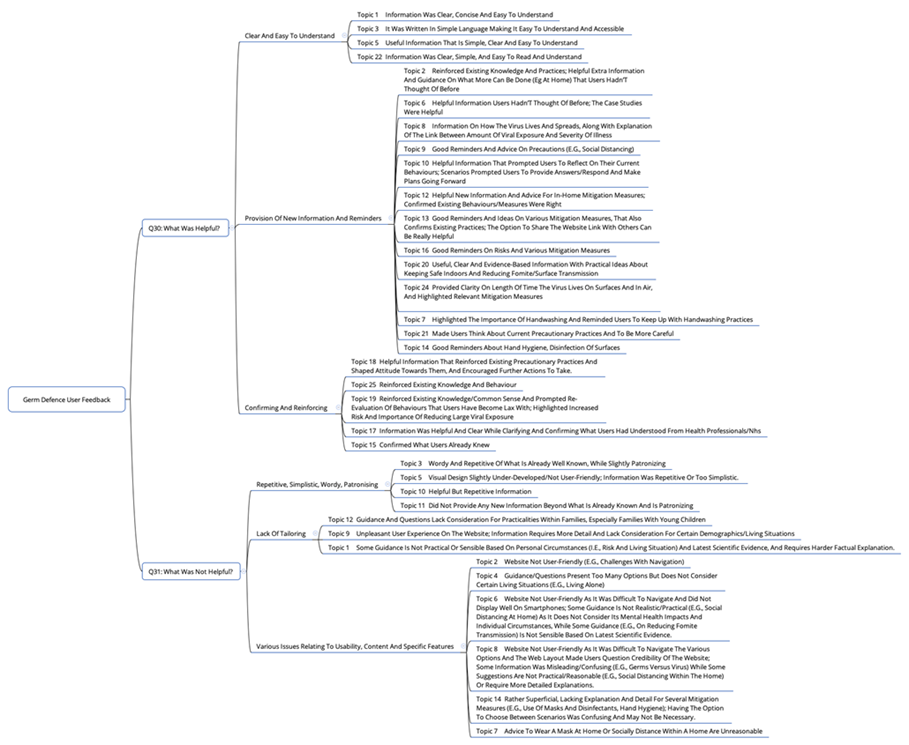
