## Supplementary material 2 for "Applying machine-learning to rapidly analyse large qualitative text datasets to inform the COVID-19 pandemic response: Comparing human and machine-assisted topic analysis techniques"

**Online supplementary material 2**

Figure 1. Diagnostic values by number of topics for the question A corpus (left) and question B corpus (right)

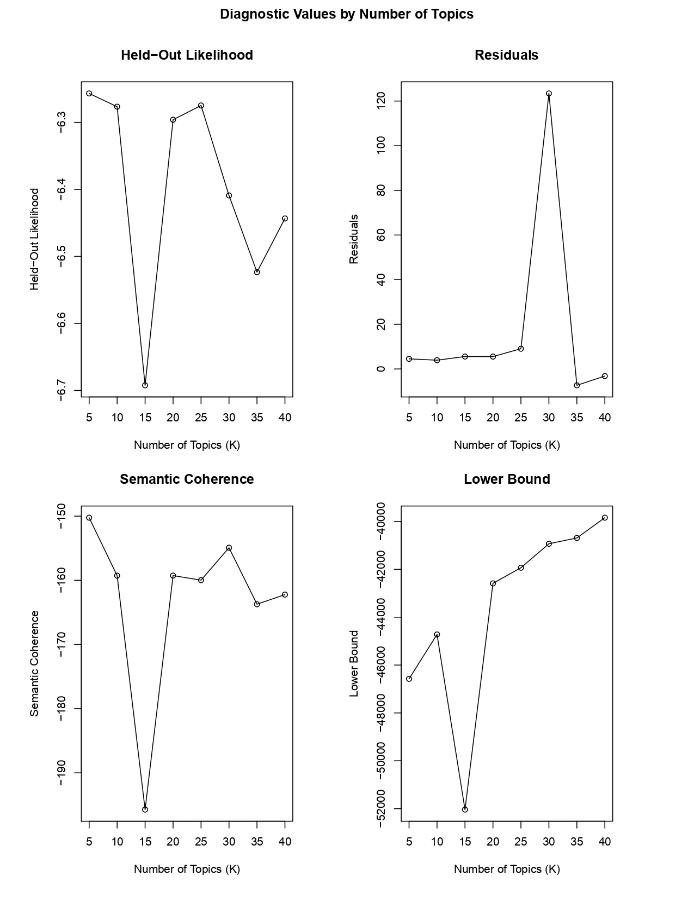

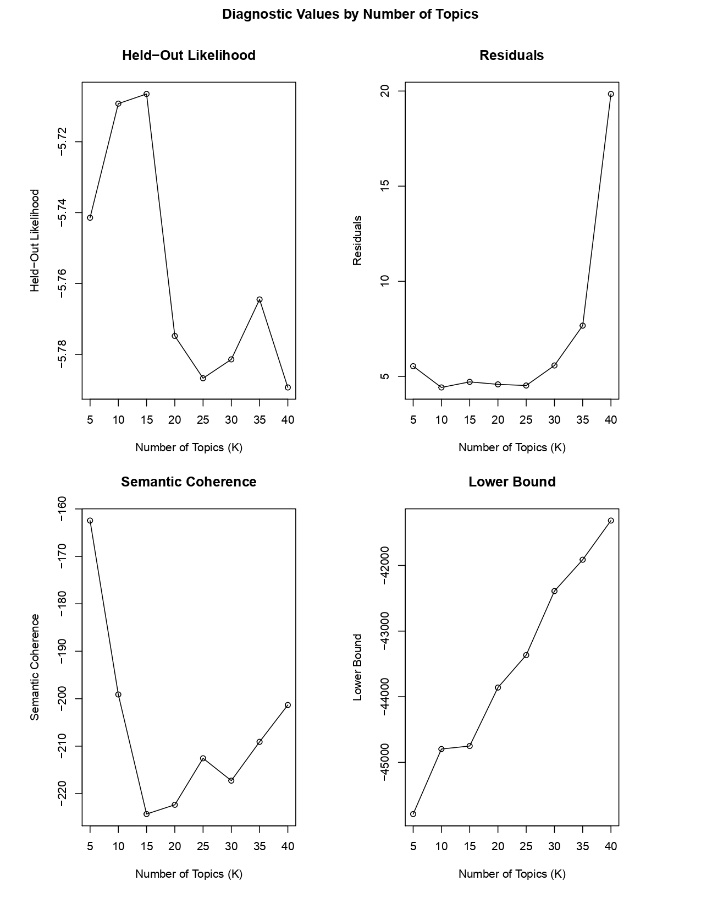
