## Supplementary material 3 for "Applying machine-learning to rapidly analyse large qualitative text datasets to inform the COVID-19 pandemic response: Comparing human and machine-assisted topic analysis techniques"

**Online supplementary material 3**

Box 1. Unsupervised machine learning output example on “What was helpful about the information on the Germ Defence website?”

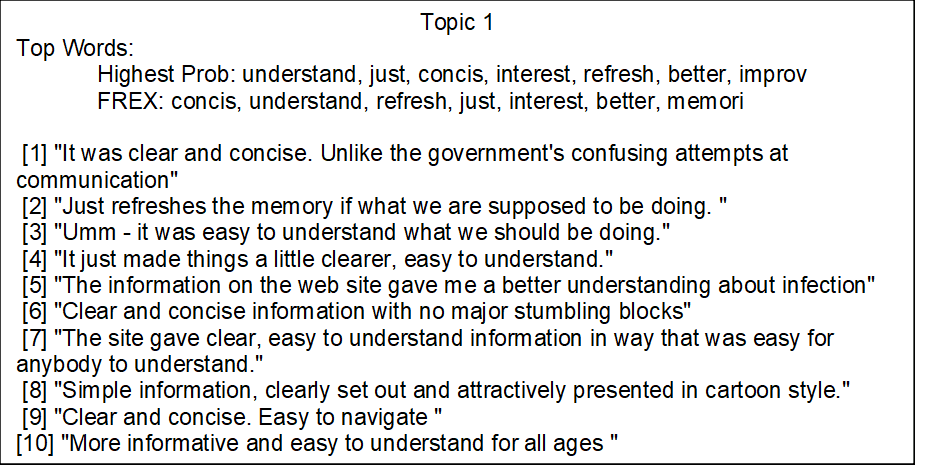

Box 2. Unsupervised machine learning output example on “What did you not find helpful about the information on the Germ Defence website?”

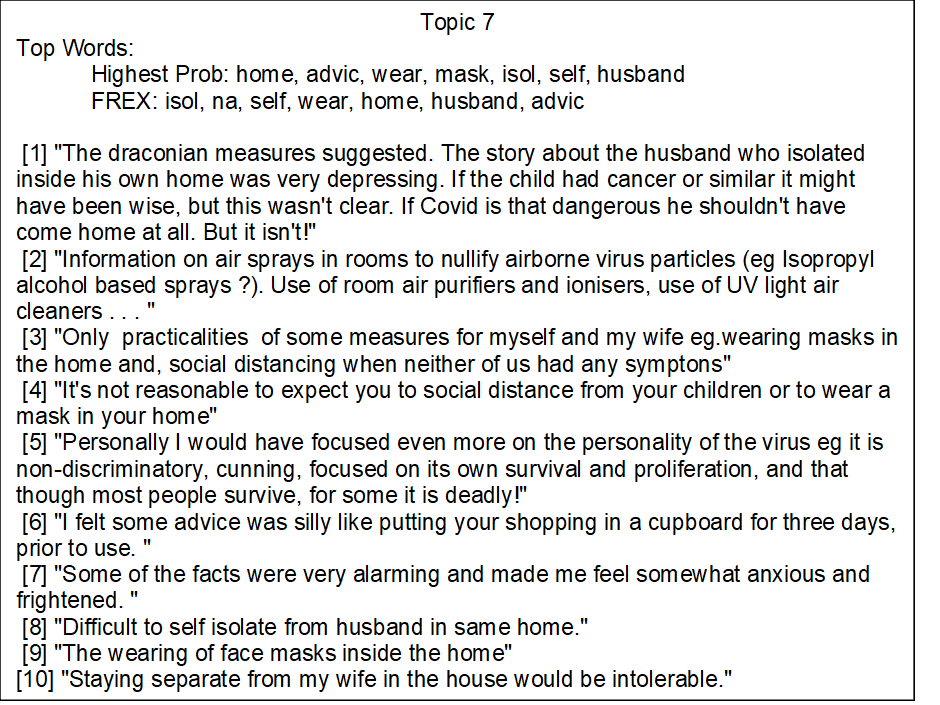
