## Supplementary material 4 for "Applying machine-learning to rapidly analyse large qualitative text datasets to inform the COVID-19 pandemic response: Comparing human and machine-assisted topic analysis techniques"

**Online supplementary material 4**

Theme descriptions and codes from the human analysis

| **Theme** | **Description** | **Codes** | **Example quotes** |
| --- | --- | --- | --- |
| **Language and layout style** | This theme encompasses opinions of how the material was presented. Many felt that the website was clear and simple, though others felt it was repetitive and lacked sophistication. | 1) Clear and simple  2) Not enough information  3) Not streamlined or sophisticated  4) Too repetitive  5) Too simplistic/patronising | “Anyone who can use a computer can follow this. It was a good easy to read set of instructions, the questions were thought-provoking.” P985  “It is 'wordy'.  There is quite a bit to get through and although the language has been made very clear, it is easy to imagine a lot of people giving up before finishing.” P1067 |
| **Confidence in how to perform the behaviours** | This website content increased confidence and self-efficacy by providing reinforcement and reminders of how to perform the behaviours, as well as practical tips and real-world examples of how to carry these out in users’ own homes. Having the information and behaviours confirmed by those perceived to be experts was reassuring and empowering. | 1) Clear practical advice and troubleshooting is helpful  2) Feeling informed and reinforced by reliable sources is empowering  3) Inconsistencies undermine confidence | “It’s the first time I have read a comprehensive and easily understandable guide relating to keeping safe in the home and which also gave readily understandable reasons and contextual relevant information.” P531  “I found the examples and actual case histories especially helpful- they made the process of infection control seem more "authentic" somehow, made the whole process sort of closer to home and made me think how I might alter my usual ways at home to be safer” P714 |
| **Reducing all or nothing thinking** | This theme encompasses the idea that small changes make a difference. Many users liked the website’s emphasis on the role of viral load, and how it encourages users to evaluate their own risk level and to act according to these evaluations. However, some still felt that there wasn’t enough balance, and the advice on Germ Defence could seem restrictive and inflexible. | 1) Trying to perform all the behaviours is exhausting  2) Understanding that small changes matter is motivating  3) We should act according to risk  4) ‘All or nothing’ thinking is demotivating  5) Some behaviours are very challenging in certain situations | “I hadn’t taken on board before that it is really beneficial to reduce the AMOUNT of COVID virus. I thought it was all or nothing! This has been helpful, especially as I’m a teacher, so was feeling a bit defeatist about the risks. It’s difficult teaching at a distance, but this makes me realise that it is worth all the measures we have in place at school.” P362  “It doesn’t feel like it realistically balances steps we can take to reduce risk with how humans actually behave. It’s like advocating for abstinence - sure, your risk of becoming pregnant is 0 if you don’t have sex. But hardly anyone is going to do that because the drive to have sex is very strong. Same with social distancing and sitting alone in a cold room with the windows open and a mask on in your own home. That will reduce the risk of catching the virus hugely, but I doubt most people would actually do it.” P328 |
