## Supplementary material 5 for "Applying machine-learning to rapidly analyse large qualitative text datasets to inform the COVID-19 pandemic response: Comparing human and machine-assisted topic analysis techniques"

**Online supplementary material 5**

Figure 1. Ranking of topics in terms of prevalence in the corpus for question A, *“What was helpful about the information on the Germ Defence website?”*
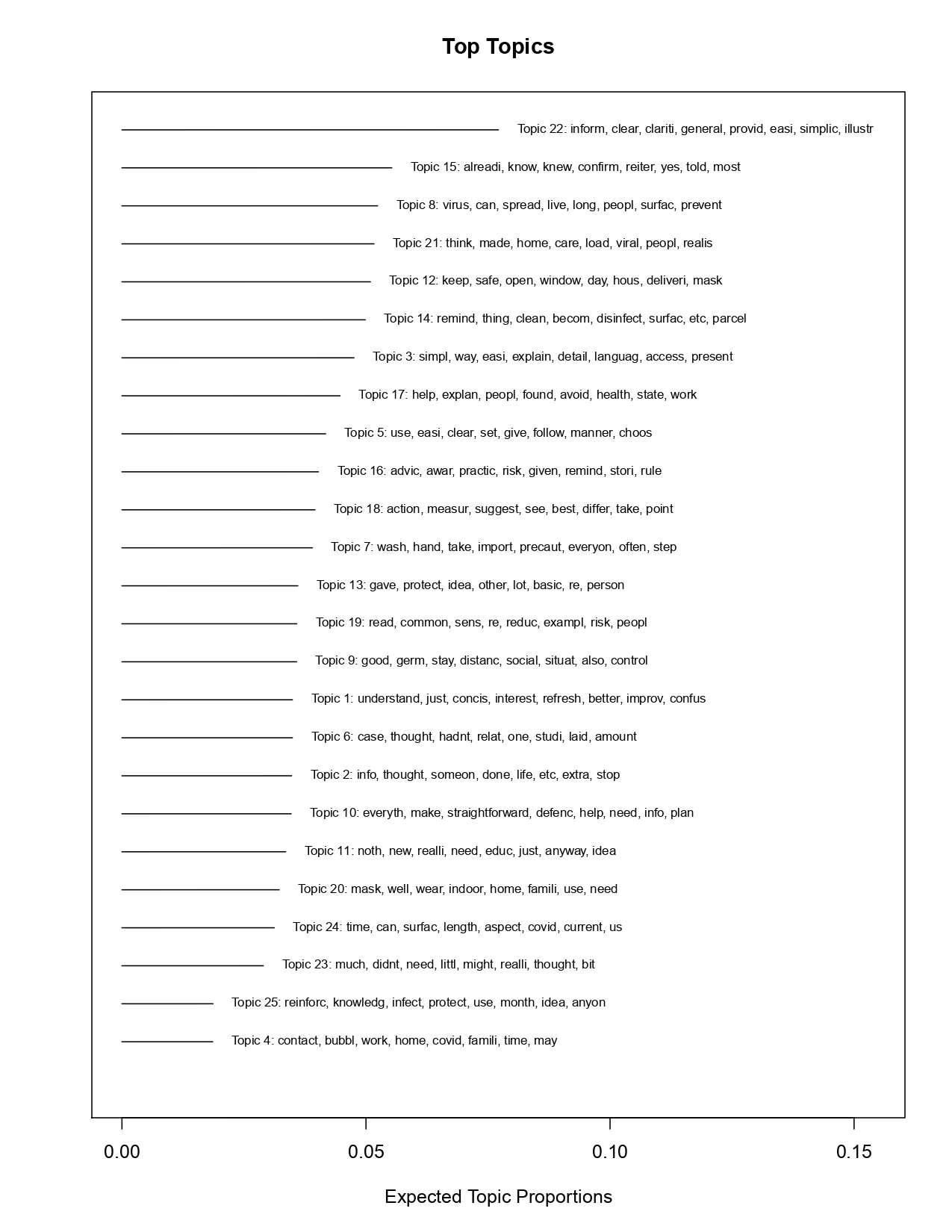

Figure 2. Ranking of topics in terms of prevalence in the corpus for question B*, “What did you not find helpful about the information on the Germ Defence website?”*

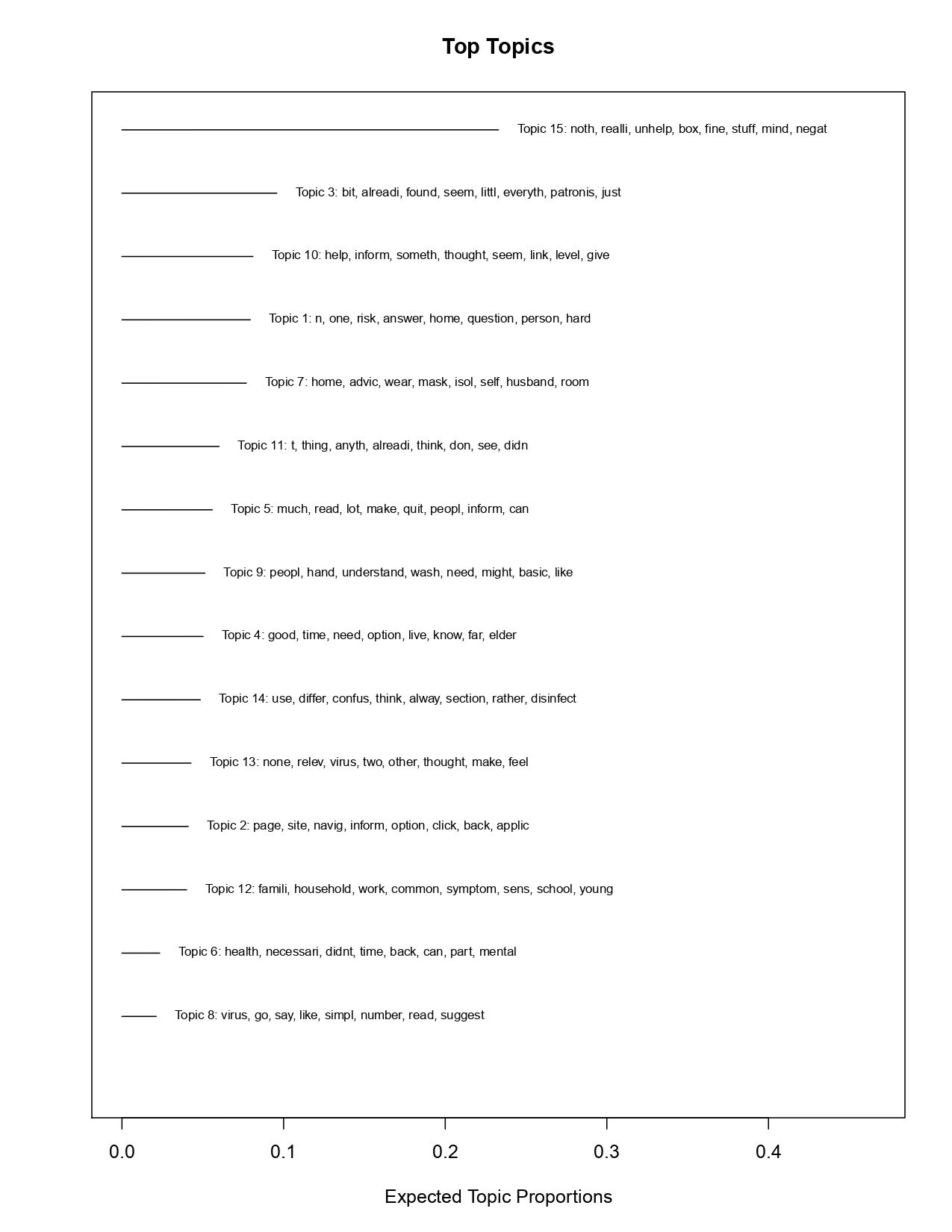
